# ANALYSIS OF A POISSON-DRIVEN STOCHASTIC COVID-19 AND HEPATITIS B CO-INFECTION MODEL

**DOI:** 10.1101/2024.08.12.24311861

**Authors:** Michael A. Pobbi, Samuel M. Naandam, Stephen E. Moore

## Abstract

In this article, we formulate and analyse a mathematical model for the co-infection of Hepatitis B virus and COVID-19. We incorporate into our framework Hepatitis B virus prevention, COVID-19 prevention, COVID-19 vaccination, and environmental factors so as to investigate their effect on transmission dynamics. First, we derive the basic reproduction number for HBV only, COVID-19 only, and co-infection stochastic models using the next generation matrix method. Next, we establish the conditions for stability in the stochastic sense for HBV only, COVID-19 only sub-models, and the co-infection model. Furthermore, we devote our attention to finding sufficient conditions for extinction and persistence. Finally, by using the Euler–Murayama scheme, we illustrate the dynamics of the co-infection, COVID-19, HBV and the effect of some parameters on disease transmission dynamics by means of numerical simulations.

## 1 Introduction

Hepatitis B disease is a life-threatening and infectious disease caused by the hepatitis B virus and is associated with serious liver infection, leading to liver malfunction, including cirrhosis, liver fibrosis, and hepatocellular carcinoma. The virus is typically transmitted from mother to child during birth and delivery, in early childhood, as well as through contact with blood, semen, or other body fluids from an infected person [61]. Between 8 weeks and 5 months, the exposed individual can experience symptoms including fever, loss of appetite, joint pain, fatigue, and nausea [12].

It remains a global health challenge, despite the development of antiviral therapies and the success of immunization [18]. Currently over 81 million people, from WHO African Region are chronically infected[3]. It is estimated that about 2 billion people are infected with HBV globally, with about 1.5 million infections annually [30], and 0.6 million annual deaths from HBV-related liver diseases [31, 6]. Countries such as Ghana, continue to have much higher (from 8.36% to 12.3%) disease prevalence rate than the global average of 3% [61] and approximately 294 HBV-related deaths weekly [13].

COVID-19, on the other hand, is caused by Severe Acute Respiratory Syndrome Corona virus 2 (SARS-CoV-2) and affects the functionality of the respiratory system. It spreads from person to person via direct contact with respiratory droplets from an infected individual. It can also be spread by touching surfaces contaminated with the virus and then touching the face. Since its emergence from Wuhan, China in December, 2019, it rapidly spread to every country and has inflicted severe public health and socio-economic burden globally [23]. It is estimated that over 7 million COVID-19 deaths and over 700 million infections have been reported since December 2019 [48]. Vaccination efforts have aided in reducing severe illness, hospitalizations, and deaths [48] worldwide. After a fourth transmission wave, Ghana also experienced a decline in COVID-19 cases. However, in July 2025, 107 confirmed cases were reported, underscoring the potential for seasonal resurgence [47].

Both COVID-19 and Hepatitis B virus (HBV) infections are life-threatening diseases and remain health problems particularly in Sub-Saharan African countries like Ghana, Botswana, and Nigeria, see, e.g., [48, 61]. Some clinical studies have confirmed the possibility of the co-infection of COVID-19 and Hepatitis B diseases, see, e.g., [63, 66, 58]. To improve understanding of the co-infection dynamics of these two global threats, mathematical models have been developed to address gaps in clinical studies and propose potential control strategies, see, e.g. [65, 56, 3, 1, 59]. In [56], the author, constructed and analysed a deterministic mathematical model that captures the co-infection dynamics of hepatitis B virus (HBV) and COVID-19 and also evaluated the impact of various intervention strategies, on the spread of both diseases in a population. In another study [59], a deterministic model was used to analyse the impact of optimal control strategies on the transmission of COVID-19 and HBV co-infection. [3] applied the Atangana-Baleanu fractional-order derivative to study a co-infection model for HBV and COVID-19. Meanwhile, [1] incorporated Gaussian noise to model the perturbations affecting the dynamics of COVID-19 and HBV co-infection. Although such studies have made significant contributions to understanding the dynamics of COVID-19 and HBV co-infection, they did not include COVID-19 vaccination, COVID-19 prevention, HBV prevention, and the combined effects of standard random fluctuations and large-scale disturbances caused by environmental factors within the same framework. This gap in the literature motivated the present work.

Several deterministic models have been developed to study the transmission dynamics of other infectious disease e.g. [15, 32, 52, 55, 29]. Stochastic models, however, provide a more nuanced understanding of disease spread, e.g., [10, 15, 41]. These epidemiological models, unlike classical deterministic counterparts, e.g. [19, 42, 54], account for the effects of random fluctuations in disease transmission dynamics, recovery rates, and the potential for co-infection by multiple diseases. To help explain the long run behaviour of disease spread, other studies such as [69, 38, 51] have presented analyses of the existence, uniqueness, and persistence of solutions to their stochastic models. In this article, we also employ suitable Lyapunov functions to develop conditions for extinction and persistence for the proposed co-infection model.

Despite advances in immunization, both HBV and COVID-19 continue to pose significant public health challenges [18, 63]. Co-infection of HBV and COVID-19 can exacerbate disease severity, leading to adverse outcomes, including increased mortality risk [66, 58]. Predicting long-term trends in the prevalence of HBV and COVID-19 co-infection, as well as identifying effective control mechanisms, is therefore essential for informed public health decision-making.

Specifically, our study seeks to investigate the following objectives;

1. To develop a compartmental model for the transmission dynamics of HBV-COVID-19 co-infection.
2. To establish the Invariance and Positivity of the Lévy driven stochastic co-infection model.
3. To determine the basic Reproductive number for deterministic and Stochastic co-infection models respectively.
4. To determine conditions for stability at Disease Free Equilibrium for deterministic and Stochastic co-infection models.

To present numerical simulations of the model.

The rest of the paper is organized as follows; Section (2) discusses the Mathematical Model and the underlying assumptions. In Section (3), we analyze the COVID-19, HBV sub-models as well as the co-infection model. Section (4) is devoted to discussing the numerical method and numerical simulation of the model. Finally, we present the conclusion in Section (5).

## 2 Mathematical Model Formulation

In this section, we consider a stochastic compartmental mathematical model that uses differential equations, see Figure (1).

### 2.1 Assumptions of the model

The following assumptions are used to formulate the model.

(*A*_1_) It is assumed that recruitment into the susceptible populations is by birth and immigration.

(*A*_2_) New human infection is through close contact with infectious humans, and contaminated environments.

(*A*_3_) It is assumed that there is homogeneous mixing in both sub-populations. (*A*_4_) COVID-19 vaccination wears off after some period of time.

(*A*_5_) The total human population *N* (*t*) is a sum of all the different compartments, i.e.

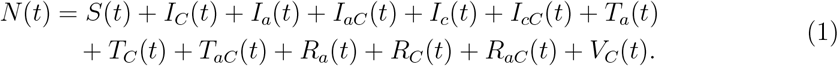

(*A*_6_) There is no relapse from HBV recovery to Susceptible state. HBV recovered recovered persons generally develop lifelong immunity and cannot be reinfected. There is, however, relapse from COVID-19 recovered state to susceptible state.

(*A*_7_) We assume that individuals with chronic HBV will remain in chronic HBV state, even if some form of medical management is administered. However they can be infected by COVID-19.

(*A*_8_) There is no vertical transmission of HBV.

### 2.2 Human Population

The model is structured into thirteen human compartments. The subpopulations of the human population at time *t*, denoted by *N* (*t*), The sub-populations of the human population at time *t*, denoted by *N* (*t*), include susceptible individuals *S*(*t*),COVID-19 infected individuals *I*_*C*_(*t*), acute hepatitis-B infected individuals *I*_*a*_(*t*), acute hepatitis-B infected and COVID-19 individuals *I*_*aC*_(*t*), chronic hepatitis-B infected individuals *I*_*c*_(*t*), chronic hepatitis-B and COVID-19 infected individ-uals *I*_*cC*_(*t*), acute hepatitis-B infected individuals receiving treatment *T*_*a*_(*t*), COVID-19 infected individuals receiving treatment *T*_*C*_(*t*), acute hepatitis-B and COVID-19 co-infected individuals receiving treatment *T*_*aC*_(*t*), acute hepatitis-B recovered individuals *R*_*a*_(*t*), COVID-19 recovered infected individuals *R*_*C*_(*t*), acute hepatitis-B and COVID-19 recovered individuals *R*_*aC*_(*t*), and COVID-19 vaccinated individuals *V*_*C*_(*t*).

The model, as seen from Figure 1, excludes HBV vaccination compartment. This is because we consider a typically high HBV prevalence population with low HBV vaccinated population (i. e., Ghana data used in estimating infections rates are typically based on such unvaccinated population). Further, HBV vaccine provides long-lasting immunity which will imply such persons can only be infected by COVID-19.

**Figure 1.**
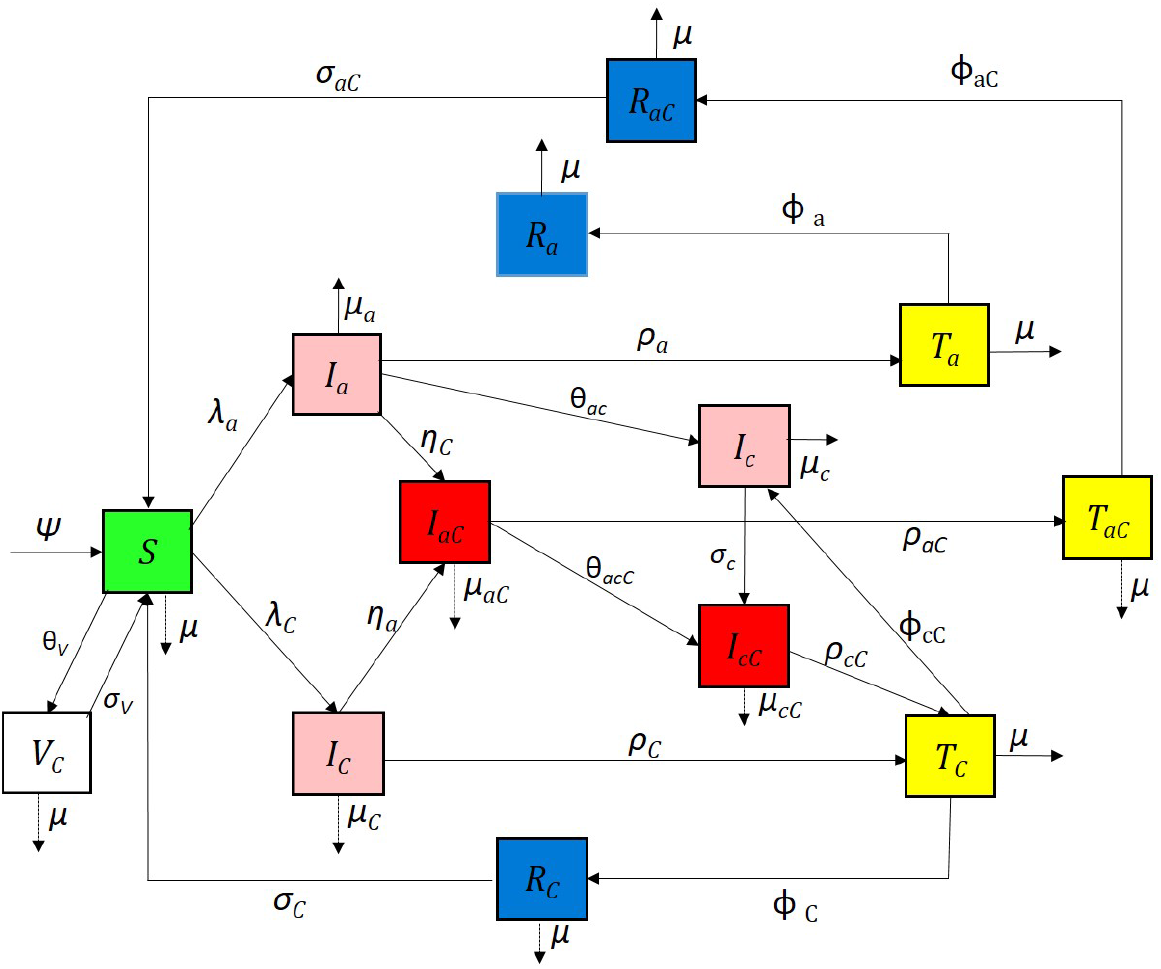
Compartmental diagram for the COVID-19 and Hepatitis B co-infection cohort.

### 2.3 Model Formulation

In this section, we consider a stochastic compartmental mathematical model that uses differential equations. Figure 1 presents the flow between states of the COVID-19 and HBV co-infection model.

We obtain the following system of non-linear differential equations from the model flow diagrams of the disease transmission shown in Figure 1.

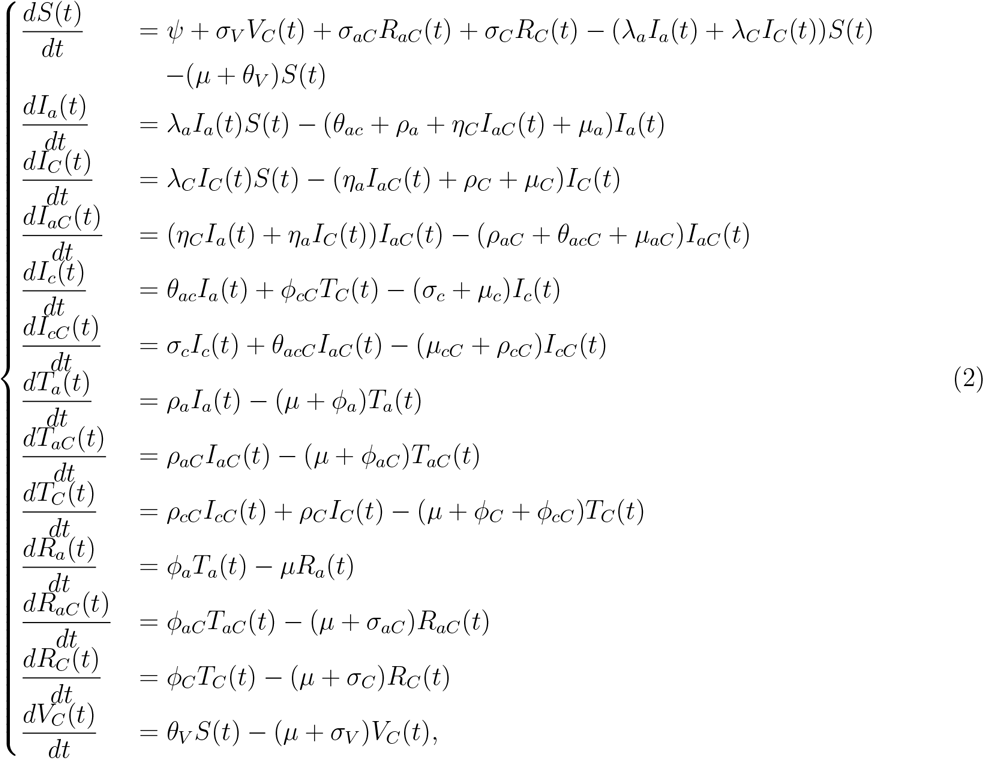

with initial conditions *S*(0) *>* 0, *I*_*C*_(0) ≥ 0, *I*_*a*_(0) ≥ 0, *I*_*aC*_(0) ≥ 0, *I*_*c*_(0) ≥ 0, *I*_*cC*_(0) ≥ 0, *T*_*a*_(0) ≥ 0, *T*_*C*_(0) ≥ 0, *T*_*aC*_(0) ≥ 0, *R*_*a*_(0) ≥ 0, *R*_*C*_(0) ≥ 0, *R*_*aC*_(0) ≥ 0, *V*_*C*_(0) ≥ 0. *λ*_*C*_, *λ*_*a*_ are the transmission rate for COVID-19, and HBV respectively. The parameters *ψ, λ*_*C*_, *η*_*C*_, *λ*_*a*_, *θ*_*aC*_, *θ*_*acC*_, *ρ*_*a*_, *ρ*_*aC*_, *ρ*_*C*_, *ϕ*_*aC*_, *ϕ*_*a*_, *ϕ*_*C*_, *σ*_*V*_, *σ*_*C*_, *σ*_*aC*_, *θ*_*V*_, *µ, µ*_*C*_, *µ*_*aC*_, *µ*_*a*_, *µ*_*c*_, and *µ*_*cC*_ are non-negative, with a forces of infection *ω*_*C*_ = *λ*_*C*_*I*_*C*_ = *β*_*C*_(1 − *κζ*)*I*_*C*_(*t*)*/N*, and *ω*_*a*_ = *λ*_*a*_*I*_*a*_ = *β*_*a*_(1− *δζ*)*I*_*a*_(*t*)*/N* respectively, and *η*_*a*_ = *ω*_*a*_, and *η*_*C*_ = *ω*_*C*_. *κ* and *δ* are fraction of the community employing protective or preventive measures for COVID-19 and HBV respectively, and *ζ* is the efficacy of personal protection strategy adopted.

The model parameters along with their description are provided in Table 1.

**Table 1:**
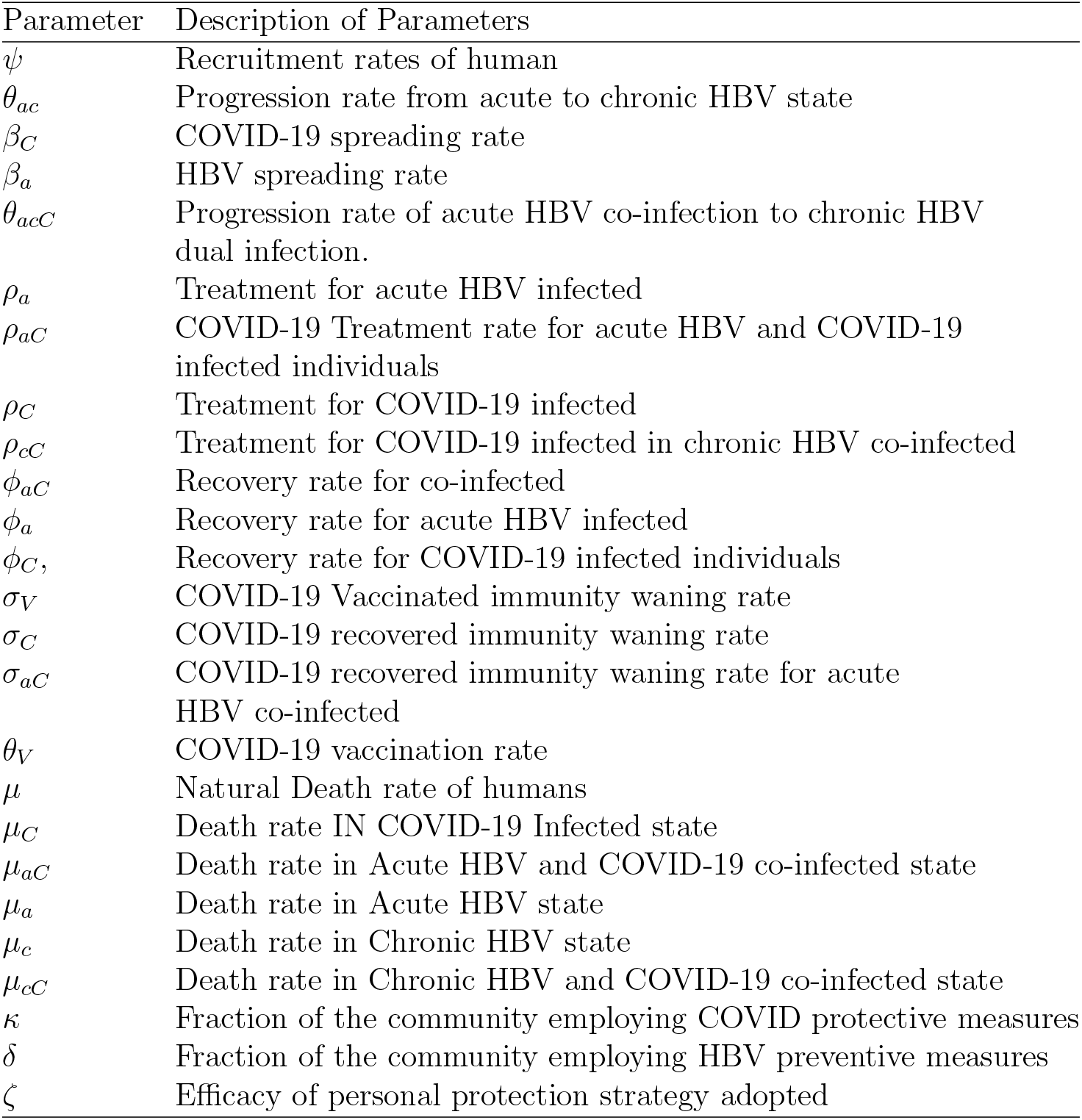
Parameters and description.

| Parameter | Description of Parameters |
| --- | --- |
| $\psi$ | Recruitment rates of human |
| $\theta_{ac}$ | Progression rate from acute to chronic HBV state |
| $\beta_C$ | COVID-19 spreading rate |
| $\beta_a$ | HBV spreading rate |
| $\theta_{acC}$ | Progression rate of acute HBV co-infection to chronic HBV dual infection. |
| $\rho_a$ | Treatment for acute HBV infected |
| $\rho_{aC}$ | COVID-19 Treatment rate for acute HBV and COVID-19 infected individuals |
| $\rho_C$ | Treatment for COVID-19 infected |
| $\rho_{cC}$ | Treatment for COVID-19 infected in chronic HBV co-infected |
| $\phi_{aC}$ | Recovery rate for co-infected |
| $\phi_a$ | Recovery rate for acute HBV infected |
| $\phi_C$ | Recovery rate for COVID-19 infected individuals |
| $\sigma_V$ | COVID-19 Vaccinated immunity waning rate |
| $\sigma_C$ | COVID-19 recovered immunity waning rate |
| $\sigma_{aC}$ | COVID-19 recovered immunity waning rate for acute HBV co-infected |
| $\theta_V$ | COVID-19 vaccination rate |
| $\mu$ | Natural Death rate of humans |
| $\mu_C$ | Death rate IN COVID-19 Infected state |
| $\mu_{aC}$ | Death rate in Acute HBV and COVID-19 co-infected state |
| $\mu_a$ | Death rate in Acute HBV state |
| $\mu_c$ | Death rate in Chronic HBV state |
| $\mu_{cC}$ | Death rate in Chronic HBV and COVID-19 co-infected state |
| $\kappa$ | Fraction of the community employing COVID protective measures |
| $\delta$ | Fraction of the community employing HBV preventive measures |
| $\zeta$ | Efficacy of personal protection strategy adopted |

Two popular ways to introduce stochastic factors into epidemic models are: (i) to assume some small and standard random fluctuation, (ii) to assume massive disturbances caused by sudden environmental shocks. The first, is described by Brownian noise while the second is described by the Lévy-driven jumps.

In this study, it is assumed that fluctuations in the environment will manifest mainly as fluctuations in the parameter *β*, i.e. *β* −→ *β* + *ϵ*_*i*_*dB*_*i*_(*t*), (*i* = 1, 2, …, 13), where *dB*_*i*_(*t*) is a one dimensional standard Brownian noise with *B*_*i*_(0) = 0, and *ϵ*_*i*_ is the intensity of the noise. Details on how this was done in the study is detailed as follows.

With regards to jumps, we restrict Lévy noise to COVID-19 infectious states. This involved the inclusion of a multiplicative jump noise which occurs randomly and proportional the number of infective in a finite population, e.g., [1]. Biological justification for the above stems from the fact that COVID-19 spreads through human interactions, and can be highly irregular due to spreader factors. Such factors, e.g., mass gatherings at funeral ceremonies and loose enforcement of public health measures (e.g., wearing of nose masks) are mentioned in Ghanaian studies, see, e.g., [46] as causes of COVID-19 jumps in the distribution of disease spread. It is observed from Figure 2 that the distribution of confirmed COVID-19 data from Ghana exhibited jumps as a result of factors outlined by [46].

**Figure 2.**
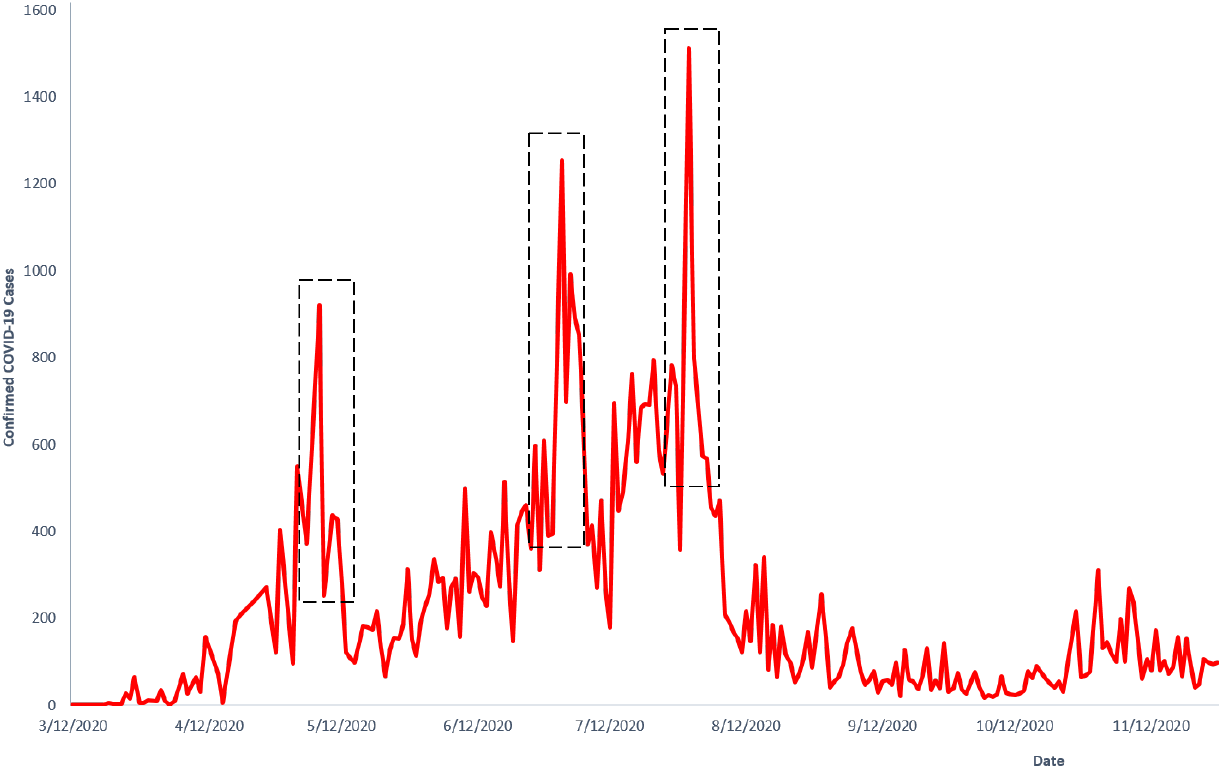
Confirmed COVID-19 cases in Ghana. [53]

Jumps are excluded in HBV. Only Brownian noise is included in the HBV infectious states as disease spread in real world data for in population considered, see, e.g.,[22] do not exhibit the presence of jumps (i.e., infection rate remains relative smooth). Hence, *I*(*t*)*dB*_*I*_(*t*) accounts for continuous fluctuations in transmission.

The Poisson-driven Lévy process, thus, is divided into linear drift term, Brownian motion and finite Lévy driven noise. The stochastic model of the system (2) takes the following form:

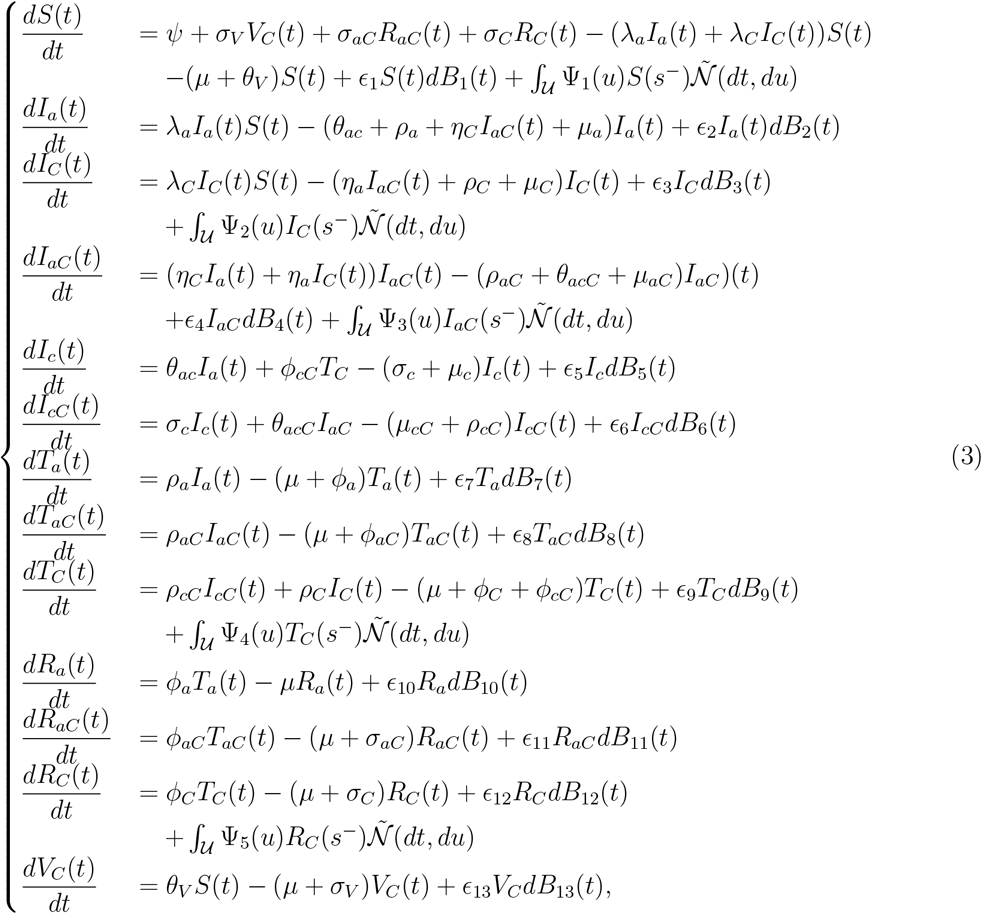

with initial conditions *S*(0) *>* 0, *I*_*C*_(0) ≥ 0, *I*_*a*_(0) ≥ 0, *I*_*aC*_(0) ≥ 0, *I*_*c*_(0) ≥ 0, *I*_*cC*_(0) ≥ 0, *T*_*a*_(0) ≥ 0, *T*_*C*_(0) ≥ 0, *T*_*aC*_(0) ≥ 0, *R*_*a*_(0) ≥ 0, *R*_*C*_(0) ≥ 0, *R*_*aC*_(0) ≥ 0, *V*_*C*_(0) ≥ 0.

The standard Brownian noise, *dB*_*i*_(*t*) is independent of 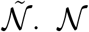 is a Poisson counting measure with compensating martingale 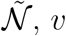 is a finite Lévy measure on a measurable subset 𝒰 ∈ (0, ∞), where *v*(𝒰) ≤ ∞. *v*(𝒰) describes the distribution of the random jump sizes in a Poisson-driven process. Again, since 𝒰 ∈ (0, ∞), then every possible jump contributes to an increase and the expectation over *v*(U) integrates over positive values, making its integral non-negative. 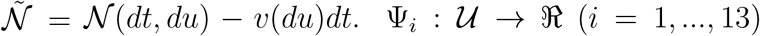 denotes the effe cts of random jumps and is bounded, and ∫_𝒰_ Ψ^2^(*X, s, u*)*v*(*du*) is bounded. We also assume that ∫_𝒰_ (1 ∧ |*x*|^2^)*ν*(*dx*) *<* ∞, thus, ensuring that the measure does not allow infinitely many jumps in a finite time interval.

We note that *S*(*t*^−^), *I*_*C*_(*t*^−^), *I*_*a*_(*t*^−^), *I*_*aC*_(*t*^−^), *I*_*c*_(*t*^−^), *I*_*cC*_(*t*^−^), *T*_*a*_(*t*^−^), *T*_*C*_(*t*^−^), *T*_*aC*_(*t*^−^), *R*_*a*_(*t*^−^), *R*_*C*_(*t*^−^), *R*_*aC*_(*t*^−^) *V*_*C*_(*t*^−^), denote the left limits of *S*(*t*), *I*_*C*_(*t*), *I*_*a*_(*t*), *I*_*aC*_(*t*), *I*_*c*_(*t*), *I*_*cC*_(*t*), *T*_*a*_(*t*), *T*_*C*_(*t*), *T*_*aC*_(*t*), *R*_*a*_(*t*), *R*_*C*_(*t*), *R*_*aC*_(*t*), *V*_*C*_(*t*), respectively.

The following definitions are necessary for the analysis of the stochastic model.

#### Definition 1

(Itô-Lévy [67]). *First, set* 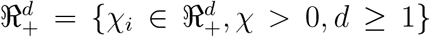. *Suppose a complete probability space* (Ω, ℱ, *P*, {ℱ_*t*_}_*t≥*0_) *with filtration* {ℱ_*t*_}_*t≥*0_, *which satisfies the usual conditions of right-continuity and completeness. B*_*i*_(*t*) *is defined on this probability space. We assume an Itô -Lévy process, X*(*t*) ∈ ℜ^+^, *of the form*

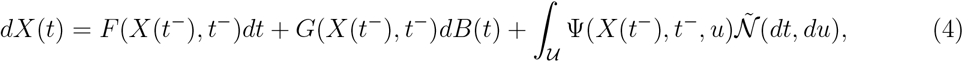

*with initial condition X(t=0) = X(0). F* : ℜ^*d*^ × ℜ^+^ × *S* → ℜ^*n*^, *G* : ℜ^*d*^ × ℜ_+_ × *S* → ℜ^*d*^, *and* Ψ : ℜ^*d*^ × ℜ_+_ × *S* × *Y* → ℜ^*d*^ *are measurable functions and X*(*t*^−^) *denotes the left limit of X*(*t*). *F* (*X*(*t*), *t*) *represents the linear drift term, G*(*X*(*t*), *t*) *the Brownian noise*, 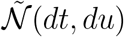 *represents the compensated Poisson random measure, and* Ψ(*X, t, u*) *represents the intensity of jumps. It is assumed that (i) v*(*U*) *is finite (i*.*e*., *the number of jumps in any finite interval is countable), (ii)* ∫_𝒰_ (1 ∧ |*x*|^2^)*ν*(*dx*) *<* ∞, *(iii)*|Ψ(*X, s, u*)| ≤ *C*(1 + |*X*|), *where C is a constant,(iv)* ∫_𝒰_ Ψ^2^(*X, s, u*)*v*(*du*) *<* ∞, *and* 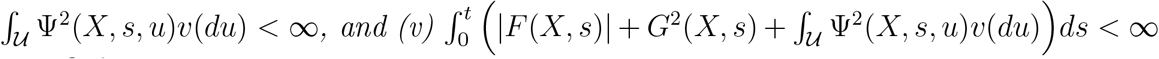 *[67] are satisfied*.

#### Definition 2

(Itô-Lévy Formula [67]). *We consider the process X expressed by* (4) *and let* 𝒱 ∈ *C*^1,2^(ℜ ^*d*^ × ℜ ^+^ × *S*; ℜ_+_) *such that* 𝒴 ≡ 𝒱 (*t, X*(*t*)), *where C*^1^ *means that the function is continuously differentiable with respect to time, and C*^2^ *means that the function has continuous second-order partial derivatives with respect to the remaining variables*.

*Then*, 𝒴 (*t*) *is again an Itô-Lévy process and*

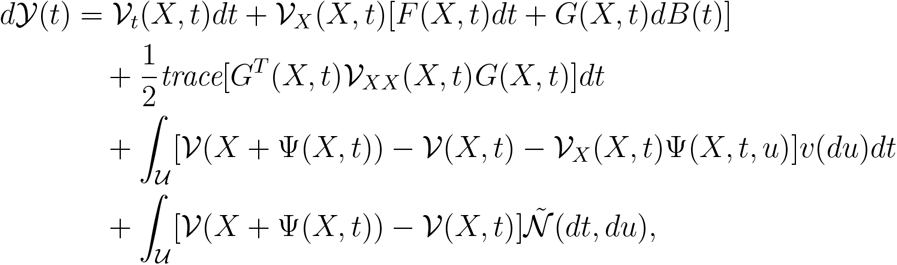

*which represents the generalized Itô’s formula with jumps where*

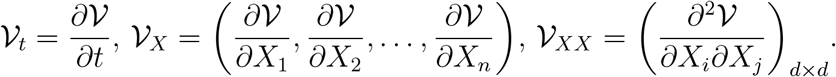

To gain insights into the underlying mechanisms that drive the spread of infectious diseases to identify the long-term behaviour of disease transmission we conduct qualitative analysis of the stochastic model (3).

## 3 Qualitative Analysis of Model

In this section, we present analysis on the existence of solution, positivity of the solution to (3), determine the reproductive number for single disease models and co-infection model, determine the conditions for Local and Global equilibrium for COVID-19 only, HBV only, and for co-infection model. Further we determine the conditions for extinction, and persistence for all three models.

### 3.1 Positivity of Model

Here, we determine the positivity of solution to the stochastic system (3) since a negative state will not be biologically meaningful. We will show the positivity of the solutions to (3). We proceed as follows

#### Theorem 1.

*Let S*(*t*^−^), *I*_*C*_(*t*^−^), *I*_*a*_(*t*^−^), *I*_*aC*_(*t*^−^), *I*_*c*_(*t*^−^), *I*_*cC*_(*t*^−^), *T*_*a*_(*t*^−^), *T*_*C*_(*t*^−^), *T*_*aC*_(*t*^−^), *R*_*a*_(*t*^−^),

*R*_*C*_(*t*^−^), *R*_*aC*_(*t*^−^) *V*_*C*_(*t*^−^), *denote the left limits of S*(*t*), *I*_*C*_(*t*), *I*_*a*_(*t*), *I*_*aC*_(*t*), *I*_*c*_(*t*), *I*_*cC*_(*t*), *T*_*a*_(*t*), *T*_*C*_(*t*), *T*_*aC*_(*t*), *R*_*a*_(*t*), *R*_*C*_(*t*), *R*_*aC*_(*t*), *V*_*C*_(*t*) *be a solution of system* (3), *given initial values* (*S*(0), *I*_*C*_(0), *I*_*a*_(0), *I*_*aC*_(0), *I*_*c*_(0), *I*_*cC*_(0), *T*_*a*_(0), *T*_*C*_(0), *T*_*aC*_(0), *R*_*a*_(0), *R*_*C*_(0), *R*_*aC*_(0), *V*_*C*_(0)) ∈ Ω, *where* Ω ∈ ℜ^+^, *then* lim_*t*→∞_ *S*(*t*) ≥ 0, lim_*t*→∞_ *I*_*C*_(*t*) ≥ 0, lim_*t*→∞_ *I*_*a*_(*t*) ≥ 0, lim_*t*→∞_ *I*_*aC*_(*t*) ≥ 0, lim_*t*→∞_ *I*_*c*_(*t*) ≥ 0, lim_*t*→∞_ *I*_*cC*_(*t*) ≥ 0, lim_*t*→∞_ *T*_*a*_(*t*) ≥ 0, lim_*t*→∞_ *T*_*C*_(*t*) ≥ 0, lim_*t*→∞_ *T*_*aC*_(*t*) ≥ 0, lim_*t*→∞_ *R*_*a*_(*t*) ≥ 0, lim_*t*→∞_ *R*_*C*_(*t*) ≥ 0, lim_*t*→∞_ *R*_*aC*_(*t*) ≥ 0, lim_*t*→∞_ *V*_*C*_(*t*) ≥ 0.

*Proof*. We determine the positivity of the stochastic system (3). We proceed to show the positivity of the solution *S*(*t*) to (3) as follows. We consider the first equation of (3),

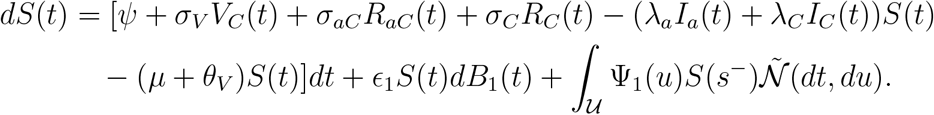

Eliminating first four terms on the right hand side and rearranging terms, we get

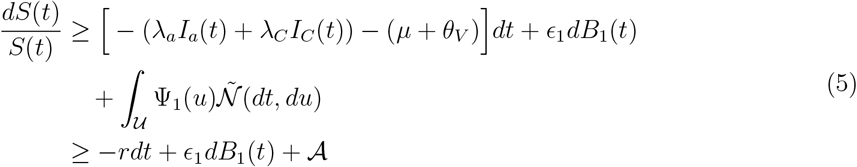

where *r* = ((*λ*_*a*_*I*_*a*_(*t*) + *λ*_*C*_*I*_*C*_(*t*) + (*µ* + *θ*_*V*_)), and 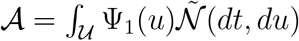. Next, we take a function *g*(*t, S*(*t*)) = ln(*t, S*(*t*)) and apply Itô-Lévy formula in (18) to get

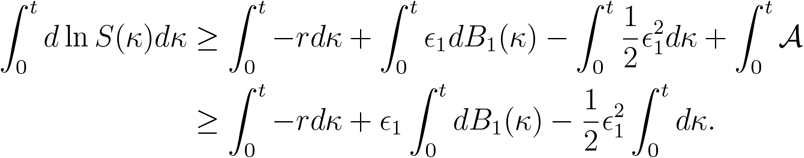

Since 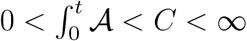, where C is a positive constant, we have

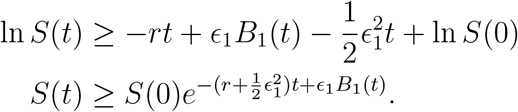

By taking the lim_*t*→∞_ on both sides we obtain

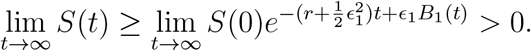

It is clear that as lim_*t*→∞_, S(t) is positive definite.

Next, we show the positivity of the solution *I*_*a*_(*t*) to (3) as follows. We consider the first equation of (3)

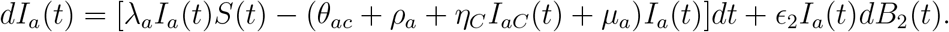

By eliminating first term on the right hand side and rearranging terms, we get

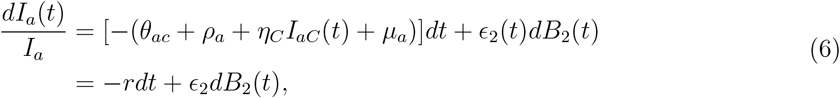

where *r* = (*θ*_*ac*_ + *ρ*_*a*_ + *η*_*C*_*I*_*aC*_(*t*) + *µ*_*a*_). Next, we take a function *g*(*t, I*_*a*_(*t*)) = ln(*t, I*_*a*_(*t*)) and apply Itô’s formula in [28] to get

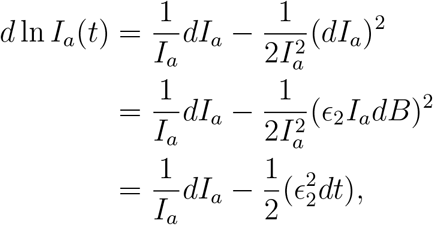

where *dB*^2^ = *dt* (by quadratic variation of Brownian motion). A rearrangement yields

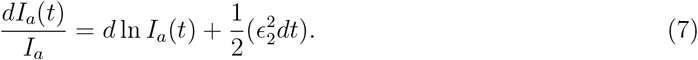

Combining (6) and (7), integrating both sides and rearrangements gives

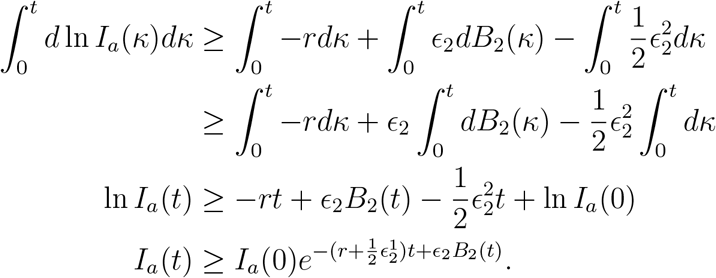

Taking the lim_*t*→∞_ on both sides we obtain

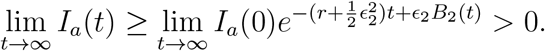

Hence *I*_*a*_(*t*) is positive definite.

We can show using similar approach that, lim_*t*→∞_ *I*_*C*_(*t*) *>* 0, lim_*t*→∞_ *I*_*aC*_(*t*) *>* 0, lim_*t*→∞_ *I*_*c*_(*t*) *>* 0, lim_*t*→∞_ *I*_*cC*_(*t*) *>* 0,lim_*t*→∞_ *T*_*a*_(*t*) *>* 0, lim_*t*→∞_ *T*_*C*_(*t*) *>* 0, lim_*t*→∞_ *T*_*aC*_(*t*) *>* 0, lim_*t*→∞_ *R*_*a*_(*t*) *>* 0,lim_*t*→∞_ *R*_*C*_(*t*) *>* 0, lim_*t*→∞_ *R*_*aC*_(*t*) *>* 0, lim_*t*→∞_ *V*_*C*_(*t*) *>* 0.

### 3.2 Invariant Region of Stochastic COVID-19 and Hepatitis B coinfection Model

An investigation of the invariant region of the model will help us determine the set of states for which the disease will eventually either persist or die out infinitely.

#### Theorem 2.

*Let N* (*t*) = *S*(*t*)+*I*_*C*_(*t*)+*I*_*a*_(*t*)+*I*_*aC*_(*t*)+*I*_*c*_(*t*)+*I*_*cC*_(*t*)+*T*_*a*_(*t*)+*T*_*C*_(*t*)+*T*_*aC*_(*t*)+*R*_*a*_(*t*)+ *R*_*C*_(*t*)+*R*_*aC*_(*t*)+*V*_*C*_(*t*) *be the population of system* (3), *then N* (*t*) *is invariant over time, that is* 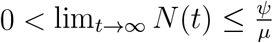, *and* (*S*(0), *I*_*C*_(0), *I*_*a*_(0), *I*_*aC*_(0), *I*_*c*_(0), *I*_*cC*_(0), *T*_*a*_(0), *T*_*C*_(0), *T*_*aC*_(0), *R*_*a*_(0), *R*_*C*_(0), *R*_*aC*_(0),

*V*_*C*_(0)) ∈ Ω.

*Proof*. From (1), we know that

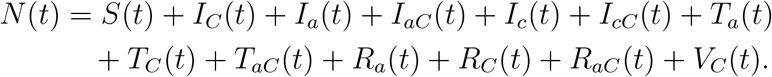

By differentiating (1) we obtain

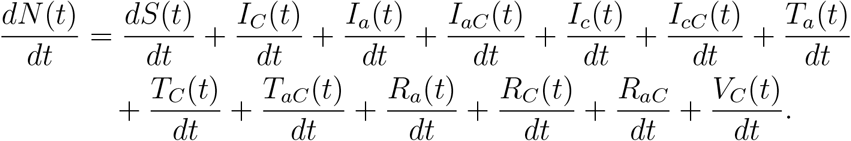

Substituting the derivatives and simplifying terms in (3) we obtain

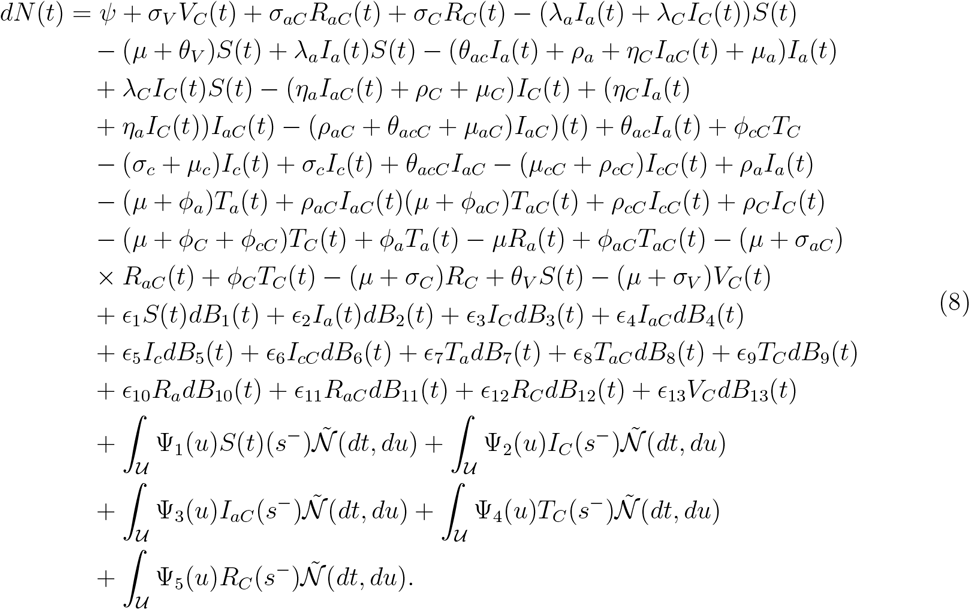

We let *µ*_*a*_ = *µ*_*aC*_ = *µ*_*c*_ = *µ*_*cC*_ = *µ*_*C*_ = *µ* and choose *ϵ* = max(*ϵ*_1_, *ϵ*_2_, *ϵ*_3_, *ϵ*_4_, *ϵ*_5_, *ϵ*_6_, *ϵ*_7_, *ϵ*_8_, *ϵ*_9_, *ϵ*_10_, *ϵ*_11_, *ϵ*_12_, *ϵ*_13_) and (∇_1_, ∇_2_, ∇_3_, ∇_4_, ∇_5_) = (*S, I*_*C*_, *I*_*aC*_, *T*_*C*_, *R*_*C*_) to obtain

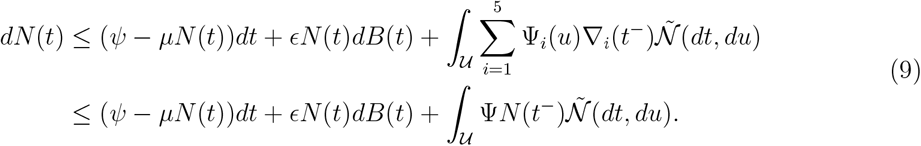

with *N* (0) *>* 0. (9) satisfies the Itô-Lévy process. By rearrangement of (9) we obtain a non-homogeneous differential equation. Next, we determine the homogeneous and particular solution and obtain the general solution to equation (9) as

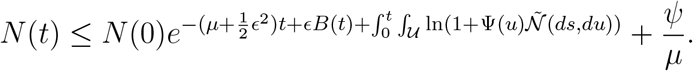

By taking the limit, we finally obtain the solution to (9) as,

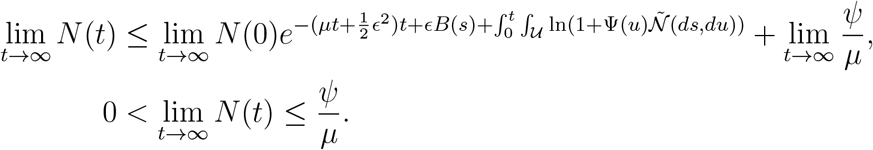

### 3.3 Disease Free Equilibrium Point

We consider the first equation of (3). For disease free equilibrium, we have *I*_*a*_(*t*) = 0, *I*_*C*_(*t*) = 0, *I*_*aC*_(*t*) = 0, *I*_*CC*_(*t*) = 0, *T*_*C*_(*t*) = 0, *T*_*a*_(*t*), *R*_*C*_(*t*) = 0, *R*_*a*_(*t*) = 0, *R*_*aC*_(*t*) = 0, *V* (*t*) = 0. We let 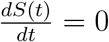,

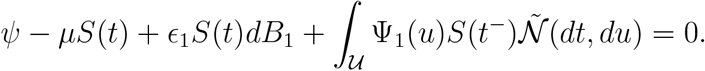

Next, we solve for *S*(*t*) as

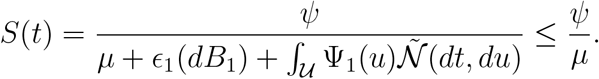

We find the expectation 𝔼 (*S*(*t*)) to get

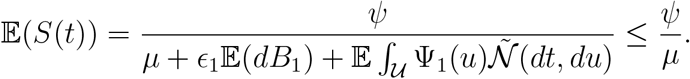

The above result is obtained by applying Definition 1 and comparison [51]. Hence, the disease free equilibrium is given by

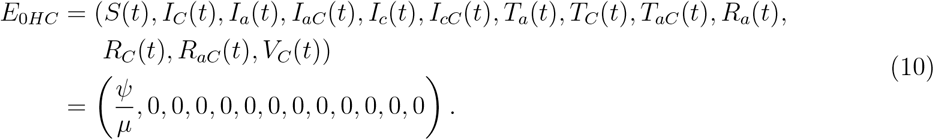

### 3.4 Reproductive Number for COVID-19 only Model

The reproduction number, ℛ_0_ of a disease determines the average number of secondary infections when an infectious person is introduced into in a completely susceptible population. It is therefore an important parameter in disease control. In this section we present results on the Reproductive Number for the deterministic COVID-19 model, 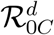, and the reproductive number for the COVID-19 model in the stochastic sense 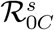. Next we determine stability conditions for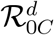, and 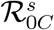 respectively.

#### 3.4.1 Basic Reproductive Number of Deterministic COVID-19 Only Model

To obtain the basic reproduction number, we consider the infected class of COVID-19 deterministic model given by

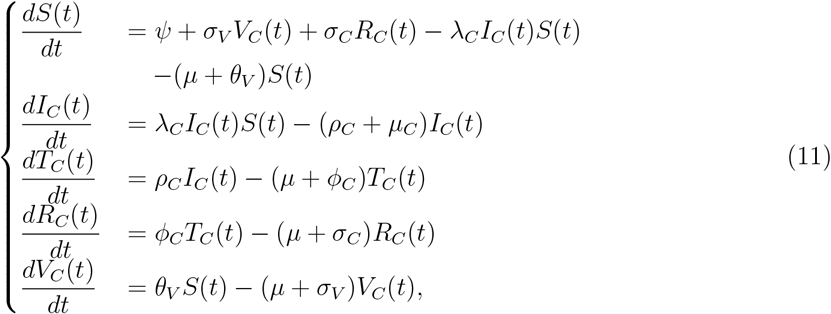

with initial conditions *S*(0) *>* 0, *I*_*C*_(0) ≥ 0, *T*_*C*_(0) ≥ 0, *R*_*C*_(0) ≥ 0, *V*_*C*_(0) ≥ 0. We employ the Next Generation Matrix (NGM) method, see, e.g., [9] to determine the basic reproductive number of the deterministic model. When using the NGM method, the basic reproductive number is defined as the spectra radius (largest eigenvalue) of *FV* ^−^1, where *F* and *V* are the partial derivatives of *f*, and *V* respectively, and *f* and *v* are the matrices of initial infections and secondary infections respectively. The inverse of *V, V* ^−1^ is defined as

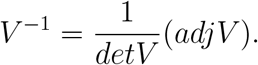

We therefore proceed by considering the primary or initial infections *f* and secondary infections *v* of infectious states of (11). The primary infections *f* is given as

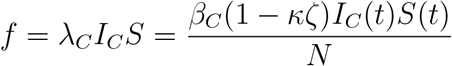

and the secondary infections *v* is given by

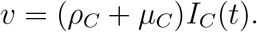

Finding the partial derivative of *f* and *v* with respect to *I*_*C*_(*t*) and evaluating at DFE we get

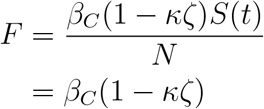

and

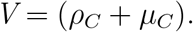

Multiplying *F* and *V* ^−1^, we get

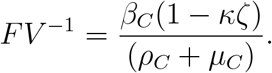

Taking the spectral radius of *FV* ^−1^, i.e., *ρ*(*FV* ^−1^), we obtain

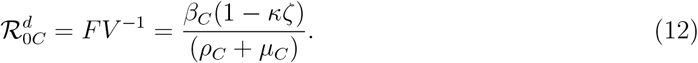

Hence the deterministic reproductive number of COVID-19 only transmission is 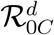.

#### 3.4.2 Basic Reproductive Number of Stochastic COVID-19 Only Model

Next, we determine the reproduction number of the COVID-19 stochastic model. We recall to the COVID-19 infected class of (3).

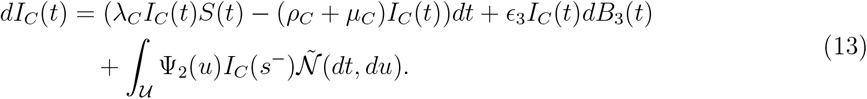

We set a function *g*(*t, I*_*C*_(*t*)) = ln(*t, I*_*C*_(*t*)) and apply Itô-Lévy formula, and evaluate at DFE to get

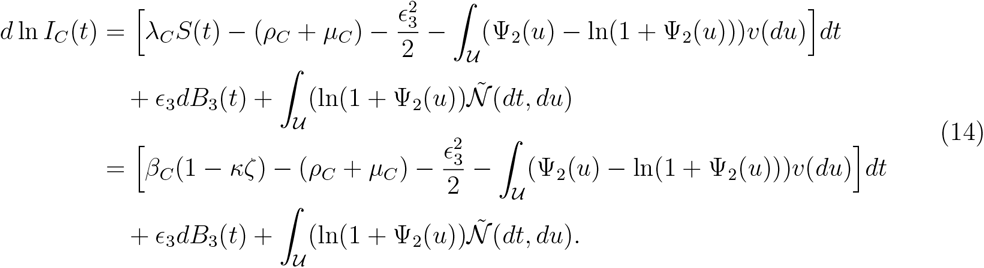

Considering partial derivatives of initial infections *F* and secondary infections *V*, we determine the basic reproduction number by means of the next generation matrix [9] as follows

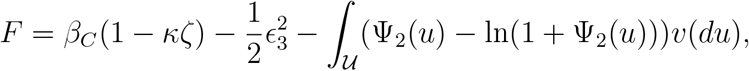

and

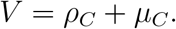

By multiplying *F* and *V* ^−1^ and factorising (*ρ*_*C*_ + *µ*_*C*_) we obtain

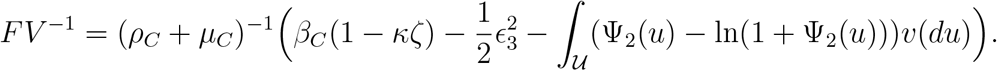

Taking the spectral radius of *FV* ^−1^, i.e., *ρ*(*FV* ^−1^), we obtain

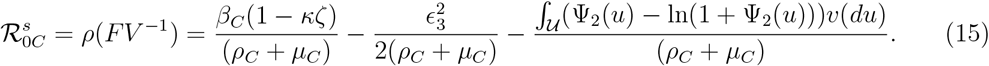

Next we examine the local stability of the 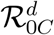 at DFE.

#### 3.4.3 Local Stability of Deterministic COVID-19 Only Model

##### Theorem 3.

*The COVID-19 disease free equilibrium point is locally asymptotically stable if and only if* 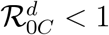 *and unstable otherwise*.

*Proof*. The Jacobian matrix of model (11) is given as

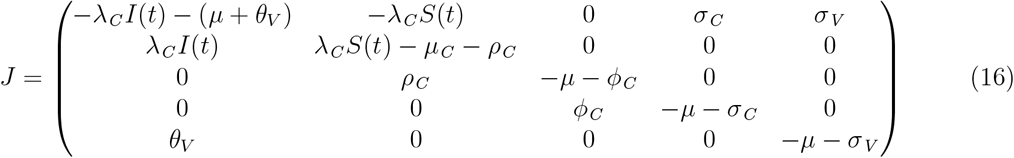

We solve (31) at the disease free equilibrium point and obtain

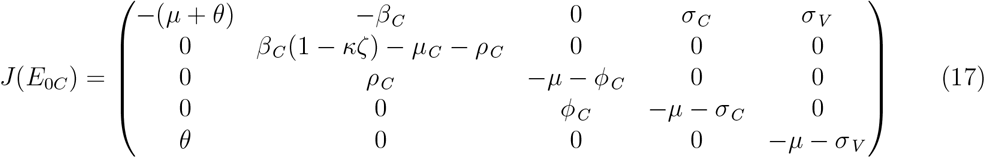

Next, we determine the characteristic matrix by det(*J*(*E*_0*C*_) − *λI*) where I is the identity matrix. The corresponding characteristic equation is given as

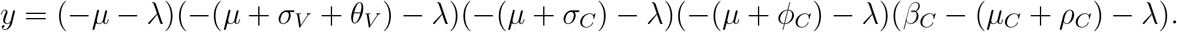

By letting det(*J*−*λI*) = 0, we observe that the three eigenvalues *λ*_1_ = −*µ, λ*_2_ = −(*µ*+*σ*_*V*_ +*θ*_*V*_), *λ*_3_ = − (*µ* + *σ*_*C*_), *λ*_4_ = − (*µ* + *ϕ*_*C*_) are negative, since all parameters are non-negative. However, for stability, all real parts of the eigenvalues must be negative. This implies

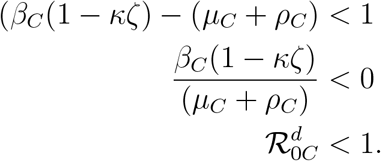

#### 3.4.4 Local Stability of Stochastic COVID-19 Only Model

It is crucial to understand the behaviour of a dynamical system near the disease-free equilibrium point. Checking for local stability at this point helps to determine whether small introductions of the COVID-19 will lead to extinction or an outbreak of the disease.

##### Theorem 4.

*If* 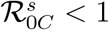, *then for any initial values of* (*S*(0), *I*_*C*_(0), *T*_*C*_(0), 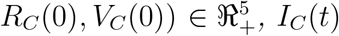 *satisfies* 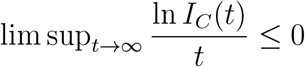.

*Proof*. We recall (18), the Itô-Lévy expression for the COVID-19 infected class of (3).

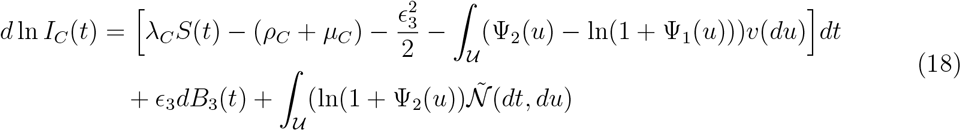

By integrating both sides of (18) and evaluating at disease free-equilibrium point, we have

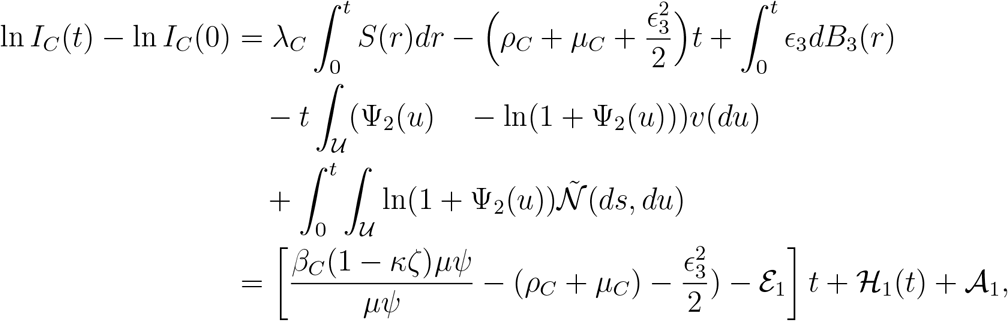

Where 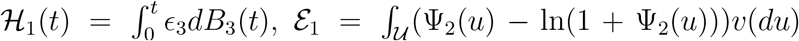, and 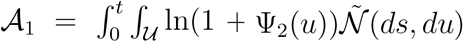 are martingales given 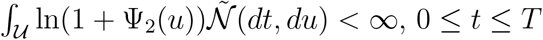.

Next, we divide through by t, take lim sup_*t*→∞_ of both sides, and apply Lemma (1) to get,

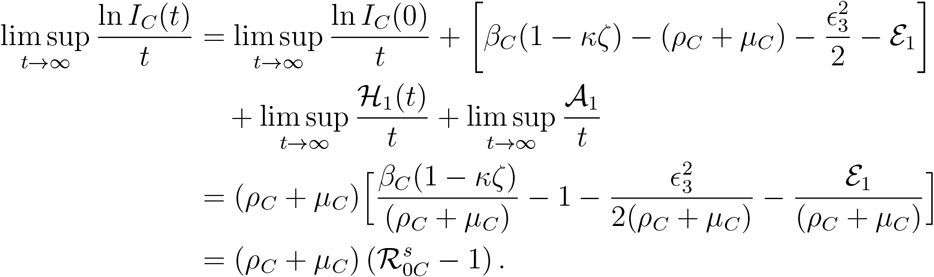

For

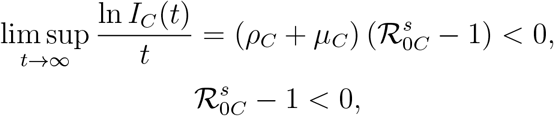

Since

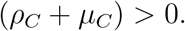

This implies that whenever 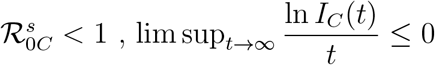.

#### 3.4.5 Global Stability of Stochastic COVID-19 Only Model

##### Theorem 5.

*If* 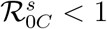, *then E*_0*C*_ *is globally asymptotically stable in* Ω.

*Proof*. We consider a Lyapunov function *L*_1_(*t*), see, e.g., [45],

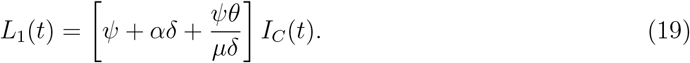

We differentiate (19), substitute (13), and evaluate at DFE to get

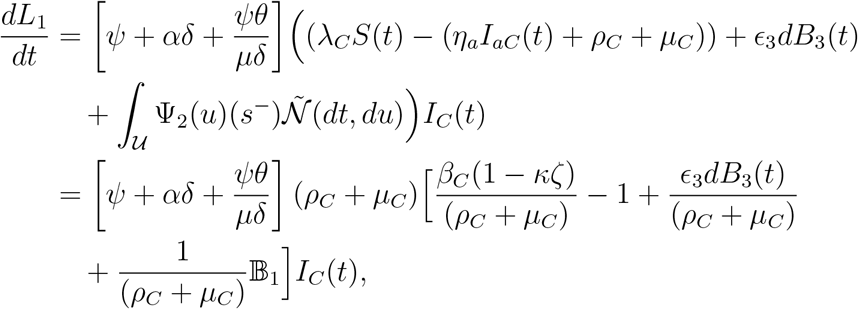

Where 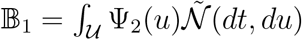, and ℰ _1_ =∫_𝒰_ (Ψ_2_(*u*) ln(1 + Ψ_2_(*u*)))*v*(*du*).

Clearly

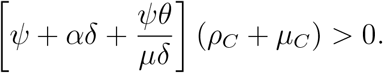

For

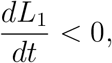

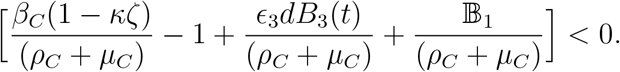

By comparison we obtain

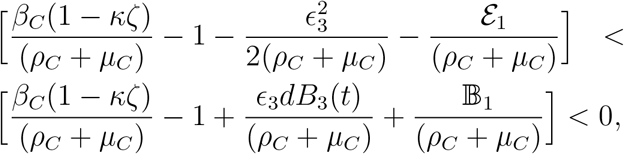

From (15) we conclude

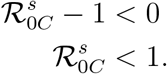

Hence, *L*_1_ is globally stable in the domain Ω.

### 3.5 Basic Reproductive Number for HBV Only Model

In this section we present results on the Reproductive Number for the deterministic reproductive number of the HBV only model, 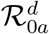, and the reproductive number of the HBV only model in the stochastic sense 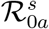. Next we determine stability conditions for 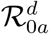, and 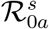 respectively.

#### 3.5.1 Basic Reproductive Number of Deterministic HBV Only Model

To obtain the basic reproduction number, we consider the infected class of HBV deterministic model given by

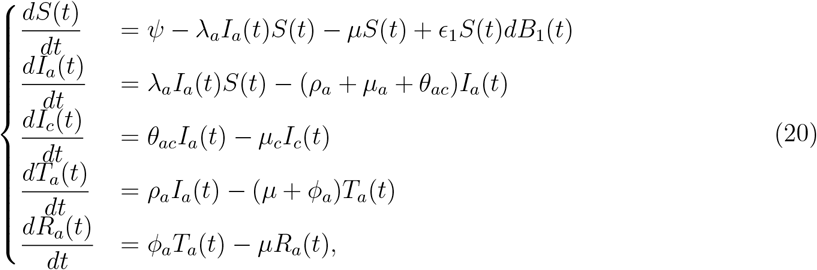

with initial conditions *S*(0) *>* 0, *I*_*a*_(0) ≥ 0, *I*_*c*_(0) ≥ 0, *T*_*a*_(0) ≥ 0, *R*_*a*_(0) ≥ 0.

We employ the next generation matrix [9] method described earlier to determine the basic reproductive number of the deterministic model. We proceed by considering the primary or initial infections *f* and secondary infections *v* of (11)

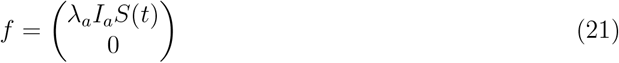

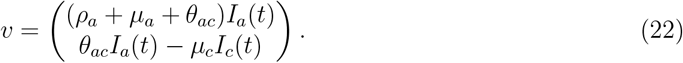

Finding the partial derivative of *f* and *v* with respect to *I*_*C*_(*t*) and evaluating at DFE gives,

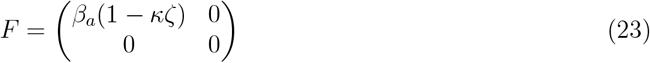

and

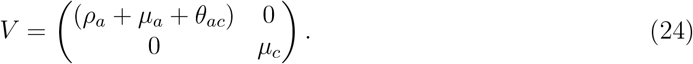

The inverse of *V* is obtained as

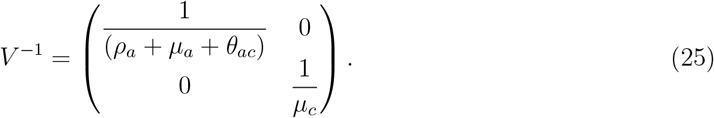

Multiplying *F* and *V* ^−1^, we get

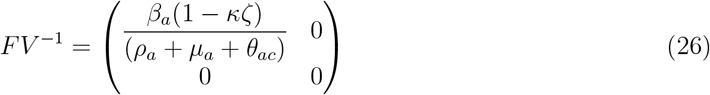

Taking the spectral radius of *FV* ^−1^, i.e., *ρ*(*FV* ^−1^), we obtain

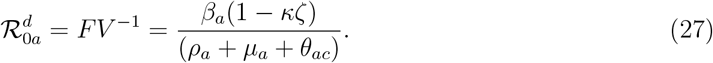

Next, we determine the reproduction number of the HBV stochastic model.

#### 3.5.2 Basic Reproductive Number for Stochastic HBV Only Model

We consider the acute HBV infectious state of the HBV only stochastic sub-model

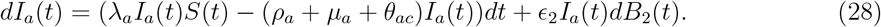

Next, we take a function *g*(*t, I*_*a*_(*t*)) = ln(*t, I*_*a*_(*t*)). Applying Itô-Lévy formula to the HBV infected class of (28), we obtain

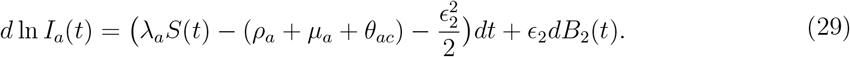

Next, we employ the next generation matrix [9] method to determine the basic reproductive number of the deterministic model. The partial derivatives with respect to *I*_*a*_ of the initial infections *F* and of secondary infections *V* are obtained and evaluated at DFE as follows

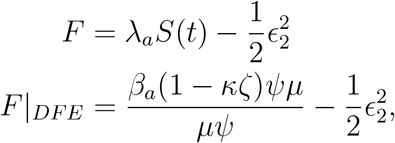

and

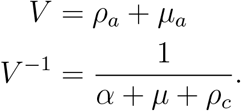

By multiplying *F* and *V* ^−1^ we get

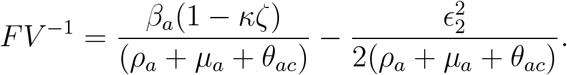

Taking the spectral radius of *FV* ^−1^, i.e., ρ(*FV* ^−1^), we obtain

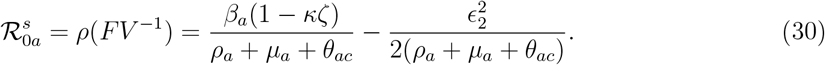

#### 3.5.3 Local Stability of Deterministic HBV Only Model

##### Theorem 6.

*The HBV disease free equilibrium point is locally asymptotically stable if and only if* 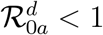 *and unstable otherwise*.

*Proof*. The Jacobian matrix of model (20) is given as

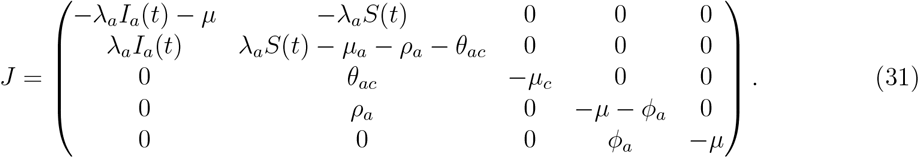

We solve (31) at the disease free equilibrium point and obtain

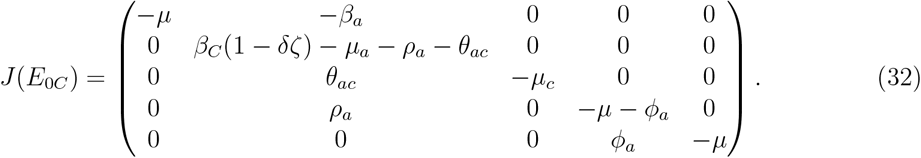

Next, we determine the characteristic matrix by det(*J*(*E*_0*C*_) − *λI*) where I is the identity matrix.

The corresponding characteristic equation is given as

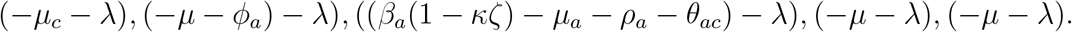

By letting det(*J* − *λI*) = 0, we observe that the three eigenvalues *λ*_1_ = −*µ*_*c*_, *λ*_2_ = −(*ϕ*_*a*_ + *µ*), *λ*_4_ = −*µ, λ*_5_ = −*µ* are negative, since all parameters are non-negative. However, for stability, all real parts of the eigenvalues must be negative. This implies

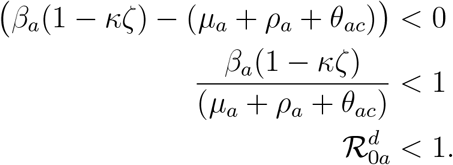

#### 3.5.4 Local Stability of Stochastic HBV Only Model

Local stability analysis at disease-free equilibrium point helps determine whether the introduction of a few infected HBV individuals in the population will become extinct or lead to an outbreak. This is crucial for understanding the behaviour of a dynamical system near that point in the presence of minor disturbances.

##### Theorem 7.

*If* 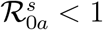, *then for any initial values of* 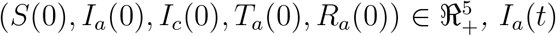 *satisfies* 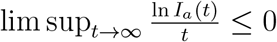.

*Proof*. We recall (28). By setting a function *g*(*t, I*_*a*_(*t*)) = ln(*t, I*_*a*_(*t*)) and applying Itô formula to (28), we obtain (29). We integrate both sides of (29) and evaluate at DFE point to obtain

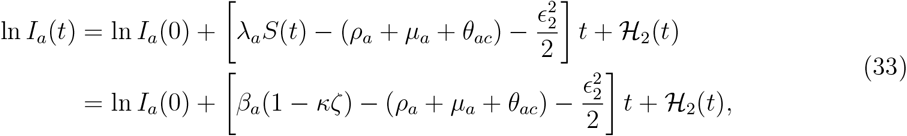

where 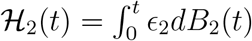 is a martingale.

Next, we divide through (33) by t, find lim sup_*t*→∞_ and use Lemma (1) to get

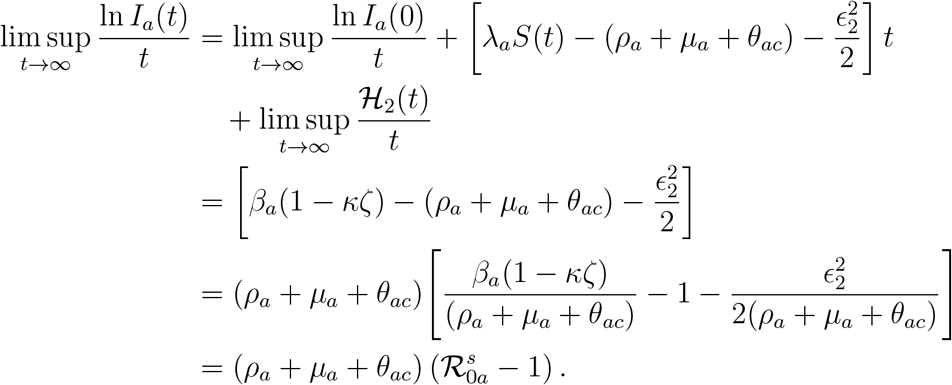

For

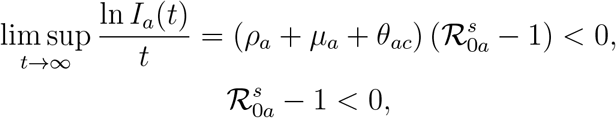

since

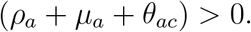

This implies that whenever 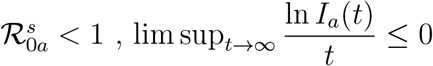.

#### 3.5.5 Global Stability of HBV Only Model

##### Theorem 8.

*If* 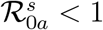, *then E*_0*a*_ *is globally asymptotically stable in* Ω.

*Proof*. We consider a Lyapunov function *L*_2_, see, e.g., [45],

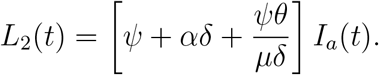

We differentiate to obtain

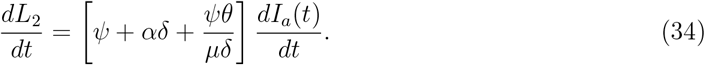

We substitute *dI*_*a*_(*t*), into (34), and then solving at DFE we get

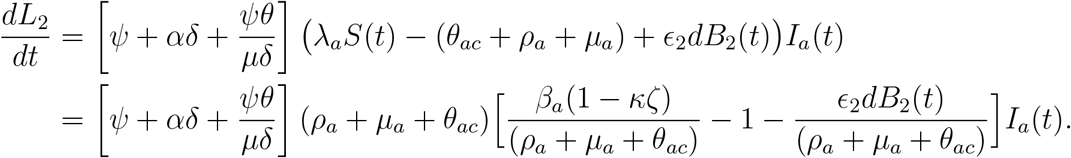

Clearly

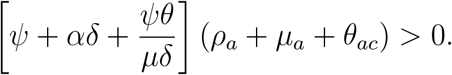

For

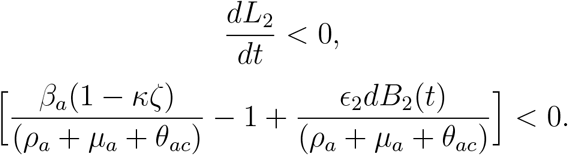

By comparison we obtain

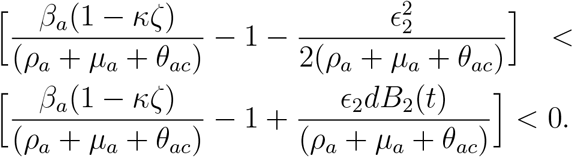

From (30) we conclude

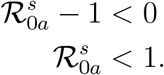

Hence *L*_2_ is globally stable in the domain Ω.

### 3.6 Reproductive Number of Co-infection Model

In this section we present results on the Reproductive Number for the deterministic co-infection model, 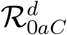, and the reproductive number for the co-infection model in the stochastic sense 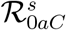. Next we determine stability conditions for 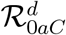, and 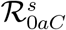 respectively.

#### 3.6.1 Reproductive Number of Deterministic Co-infection Model

In this section, we employ the next generation matrix [9] method to determine the basic reproductive number of the deterministic co-infection model. As explained earlier, the NGM method for determining the basic reproductive number requires that we first find *f*, the matrix of new infections and *v*, the matrix of secondary infections respectively. We therefore proceed by considering only disease infectious states of (2) we have

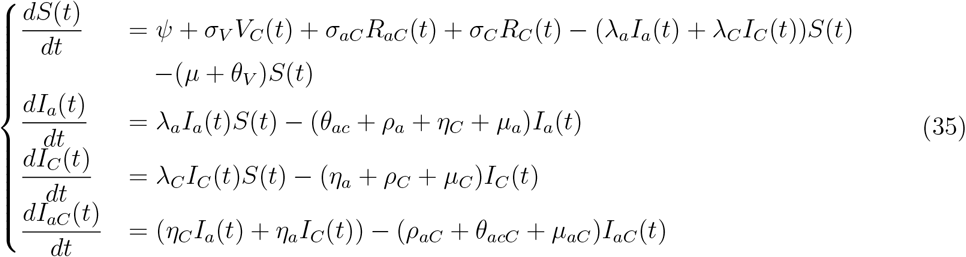

with initial conditions *S*(0) *>* 0, *I*_*C*_(0) ≥ 0, *I*_*a*_(0) ≥ 0, *I*_*aC*_(0) ≥ 0, *I*_*c*_(0) ≥ 0, *I*_*cC*_(0) ≥ 0. *λ*_*a*_*I*_*a*_*S* and *λ*_*C*_*I*_*C*_*S* represent new infections.*η*_*C*_*I*_*a*_*I*_*aC*_ and *η*_*a*_*I*_*C*_*I*_*aC*_ represent progression to co-infection. We determine *f* which represents primary or new infections from the susceptible compartment and *v* which represents the rate of transition from one compartment to the other. These are represented as

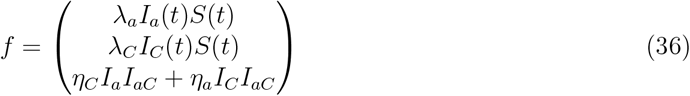

and

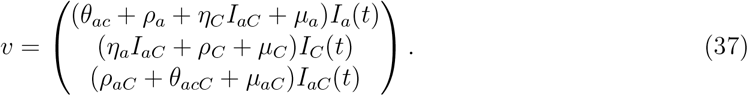

Using the NGM method we seek to determine the basic reproductive number which is the spectra radius of the *FV* ^−1^. Finding the partial derivatives *F* and *V* and evaluating at DFE gives,

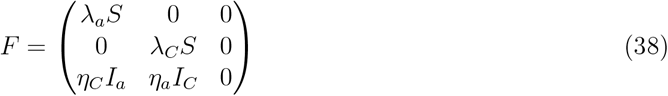

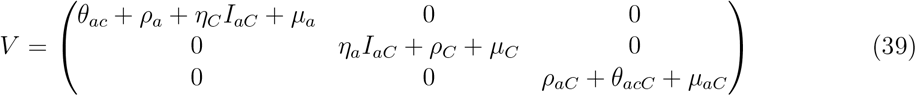

Next, we determine *FV* ^−1^, where 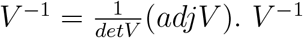 is computed as

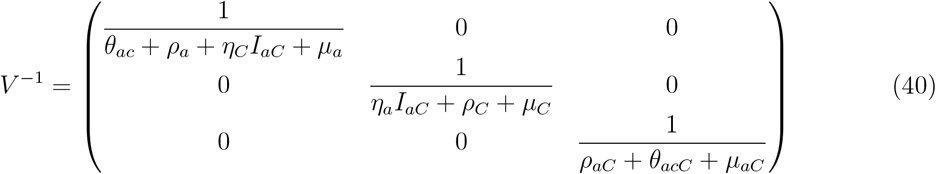

Multiplying *F* and *V* ^−1^ gives

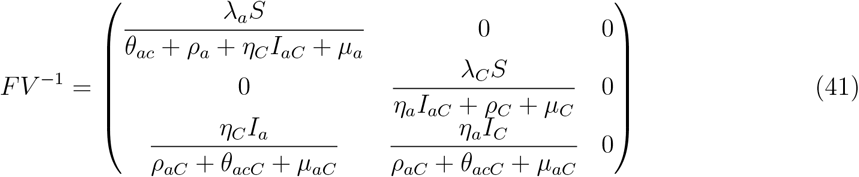

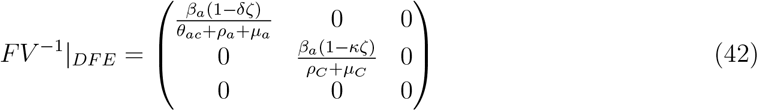

where

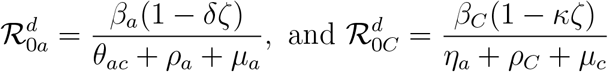

are basic reproduction numbers for the deterministic HBV and COVID-19 only models in the co-infection model (2).

The basic reproduction numbers 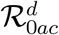 associated with (2) is given by the spectral radius (i.e. largest eigenvalue) of *FV* ^−1^. Hence, reproduction number for the full model system (2) is given by

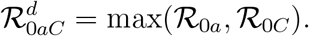

#### 3.6.2 Basic Reproductive Number of Stochastic Co-infection Model

In this section we determine the reproduction number of the COVID-19 and acute HBV stochastic co-infection model. We consider the co-infected class of (3).

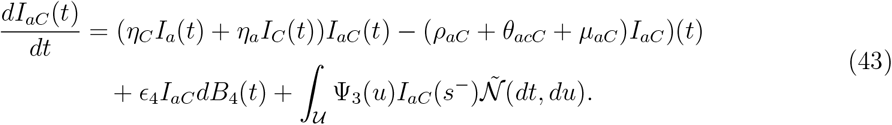

We set a function *g*(*t, I*_*C*_(*t*)) = ln(*t, I*_*C*_(*t*)). Applying Itô-Lévy formula to (43), we obtain

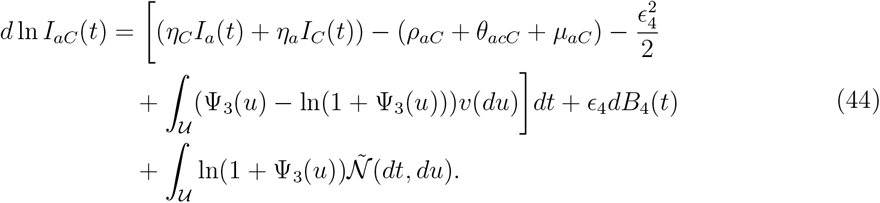

Considering partial derivatives with respect to *I*_*aC*_ of the initial infections *F* and of secondary infections *V*, we determine the basic reproduction number by means of the next generation matrix [9] as follows

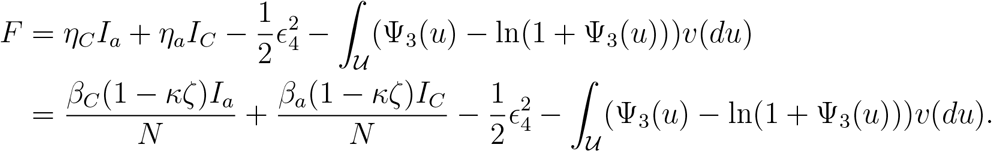

Since

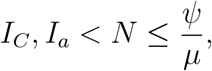

we have

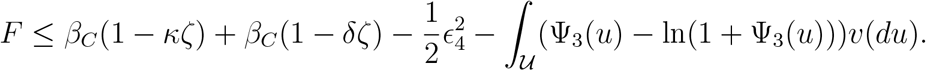

Also

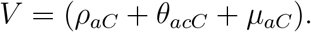

Multiplying *F* and *V* ^−1^ and factorising (*ρ*_*aC*_ + *θ*_*acC*_ + *µ*_*aC*_) we get

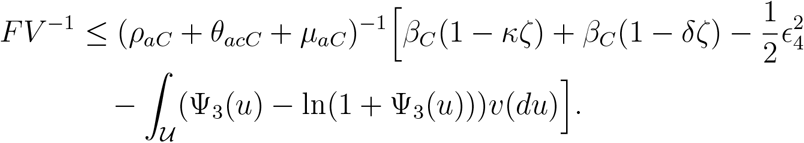

Taking the spectral radius of *FV* ^−1^, i.e. 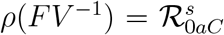, we obtain

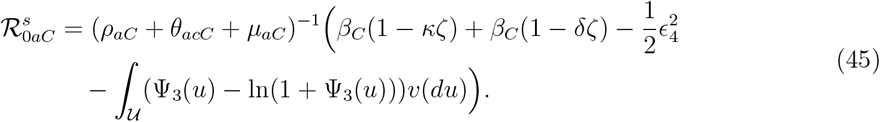

#### 3.6.3 Local Stability of Deterministic Co-infection Model

##### Theorem 9.

*The Co-infection disease free equilibrium point is locally asymptotically stable if and only if* 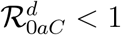 *and unstable otherwise*.

*Proof*. We recall (10). The local stability of the COVID-19-HBV co-infection disease free equilibrium is established by showing that the eigenvalues of Jacobian of the system, *J*(*Q*), are all negative and the determinant at DFE, det(*J*(*E*_0_)) *>* 0. First, we derived the Jacobian matrix of model (2) as

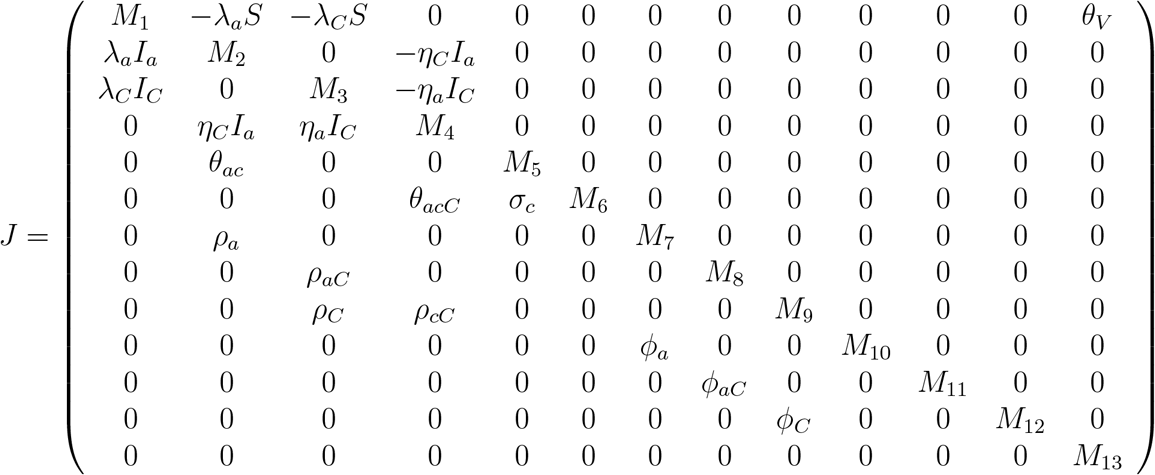

where *M*_1_ = −*λ*_*a*_*I*_*a*_ − *λ*_*C*_*I*_*C*_ − (*µ* + *θ*_*V*_), *M*_2_ = −(*θ*_*ac*_ + *ρ*_*a*_ + *η*_*C*_*I*_*aC*_ + *µ*_*a*_), *M*_3_ = −(*η*_*a*_*I*_*aC*_ + *ρ*_*C*_ + *µ*_*C*_), *M*_4_ = −(*ρ*_*aC*_ +*θ*_*acC*_ +*µ*_*aC*_), *M*_5_ = −(*σ*_*c*_ +*µ*_*c*_), *M*_6_ = −(*µ*_*cC*_ +*ρ*_*cC*_), *M*_7_ = −(*µ*+*ϕ*_*a*_), *M*_8_ = −(*µ*+*ϕ*_*aC*_), *M*_9_ = −(*µ*+*ϕ*_*C*_ +*ϕ*_*cC*_), *M*_10_ = −*µ, M*_11_ = −(*µ*+*σ*_*aC*_), *M*_12_ = −(*µ*+*σ*_*C*_), *M*_13_ = −(*µ*+*σ*_*V*_). Next, we solve *J* at the disease free equilibrium point and obtain *J*(*E*_0_). Then determine the characteristic matrix by det(*J*(*E*_0*C*_) − *λI*), where I is the identity matrix. The corresponding characteristic equation is given as

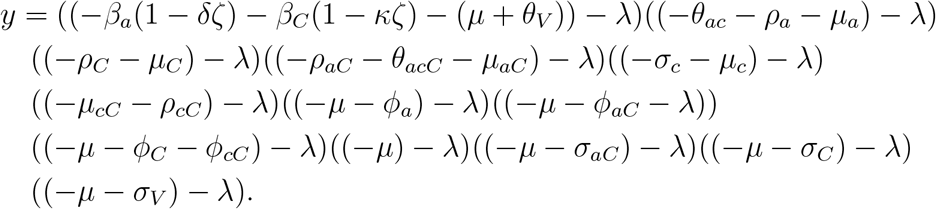

By letting det(*J*(*E*_0_) − *λI*) = 0, we observe that the thirteen eigenvalues are negative, since all parameters are non-negative. Also, since the Jacobian matrix at DFE is lower triangular, its determinant is obtained as the product of its diagonal elements as follows

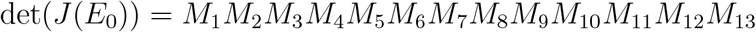

where *M*_2_, *M*_3_, …, *M*_13_ are strictly negative because they represent death rates, recovery rates, and transition rates, which are always positive. The determinant remains positive if *M*_1_ *>* 0, hence

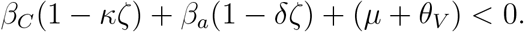

For (*ρ*_*aC*_ + *θ*_*acC*_ + *µ*_*aC*_) *<* (*µ* + *θ*_*V*_) we conclude that the DFE is locally stable when

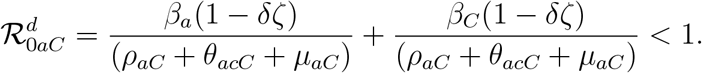

#### 3.6.4 Local Stability of Disease Free Equilibrium point in the Co-infection Model

##### Theorem 10.

*If* 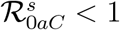, *then for any initial values of* (*S*(0), *I*_*C*_(0), *I*_*a*_(0), *I*_*aC*_(0), *I*_*c*_(0), *I*_*cC*_(0), *T*_*a*_(0*), T*_*C*_(0*), T*_*aC*_(0*),R*_*a*_*(0),R*_*C*_(0*),R*_*aC*_(0*), V*_*C*_(0)) ∈ ℜ^13^, *I*_*aC*_*(t) satisfies* 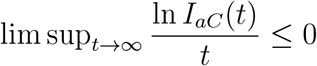.

*Proof*. We set a function *g*(*t, I*_*aC*_(*t*)) = ln(*t, I*_*aC*_(*t*)), and apply Itô-Lévy formula to the co-infected equation of (3) to obtain

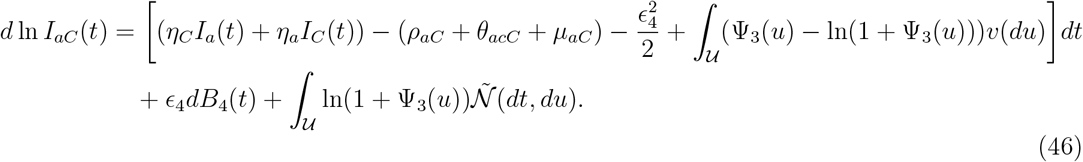

Next, we integrate both sides (46) to get

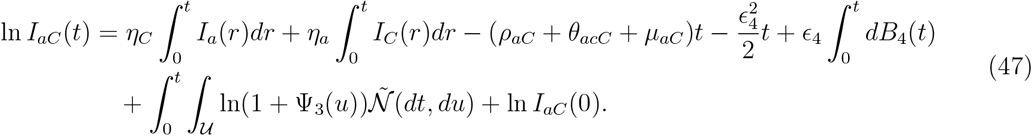

Evaluating at DFE, and since

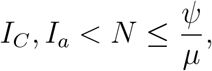

we have

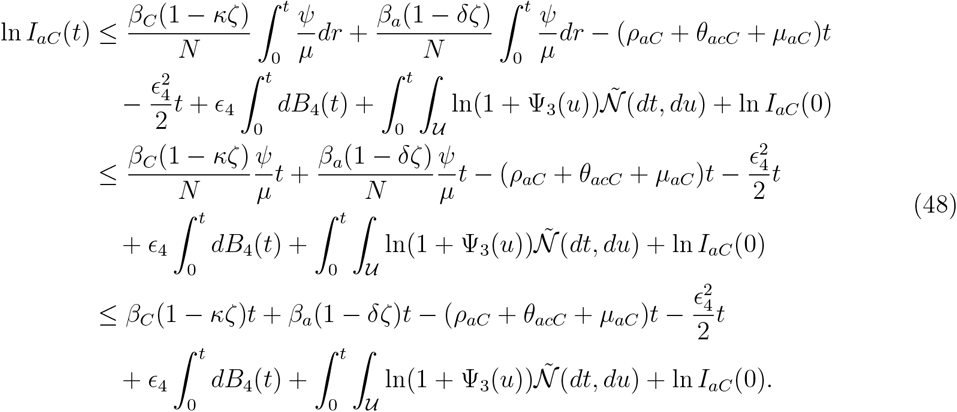

We let 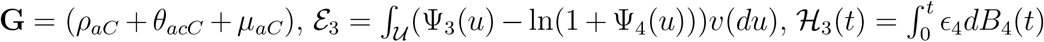, and 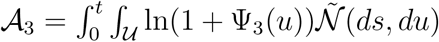. Next, we divide through (48) by t, find lim sup_*t*→∞_ and apply Lemma (1) as follows,

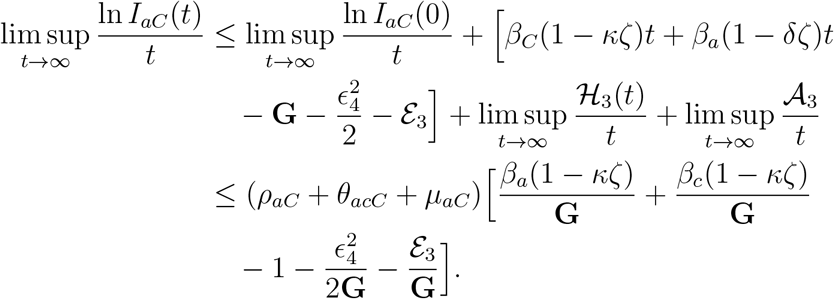

From (45) we have

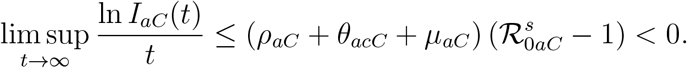

Clearly

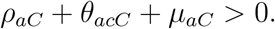

If

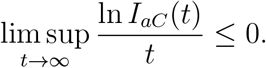

Then

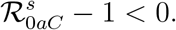

We conclude that whenever 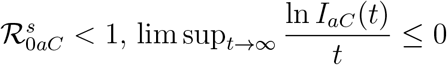.

#### 3.6.5 Global Stability of Co-infection Model

##### Theorem 11.

*If* 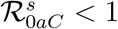, *then E*_0*aC*_ *is globally asymptotically stable in* Ω.

*Proof*. We consider a Lyapunov function *L*_3_, see, e.g., [45],

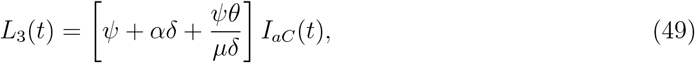

Differentiating (49) and substitution yields

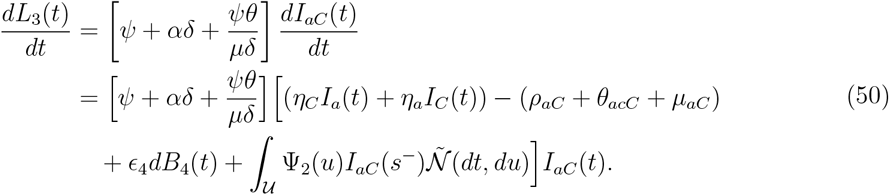

Solving (50) at DFE, and since 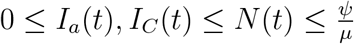 we obtain

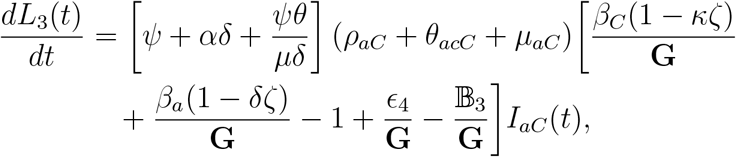

where 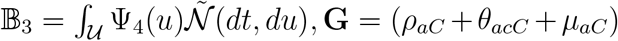, and ℰ_3_ = ∫_𝒰_ (Ψ_4_(*u*) ln(1 +Ψ_4_(*u*)))*v*(*du*).

Clearly

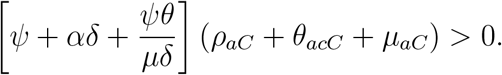

For

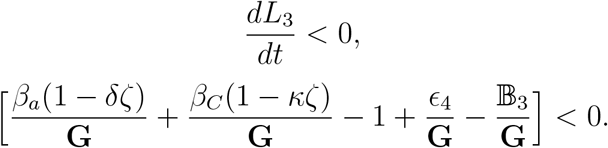

By comparison we obtain

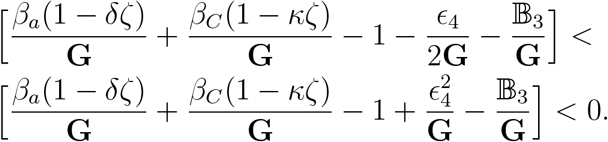

From (45) we conclude

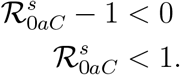

Hence *L*_3_ is globally stable in the domain Ω.

### 3.7 Extinction of HBV-COVID-19 co-infection Disease

Next, we determine conditions under which the disease will eventually die out in the population with a probability of one. In this section, we study the conditions of extinction for the co-infection model.

#### Definition 3

([1]). *We define* 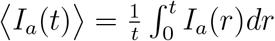, *and* 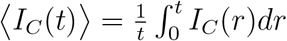.

#### Lemma 1

([67]). *Let* (*S*(*t*), *I*_*C*_(*t*), *I*_*a*_(*t*), *I*_*aC*_(*t*), *I*_*c*_(*t*), *I*_*cC*_(*t*), *T*_*a*_(*t*), *T*_*C*_(*t*), *T*_*aC*_(*t*), *R*_*a*_(*t*), *R*_*C*_(*t*), *R*_*aC*_(*t*), *V*_*C*_(*t*)) *be the positive solution of system* (4) *with given initial condition* 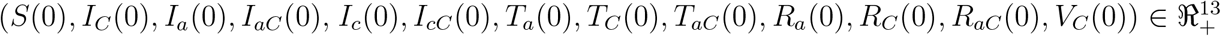. *Let also N(t) be the positive solution of equation* (4) *with given initial condition given condition N* (0) = *S*(0) + *I*_*C*_(0) + *I*_*a*_(0) + *I*_*aC*_(0) + *I*_*c*_(0) + *I*_*cC*_(0) + *T*_*a*_(0) + *T*_*C*_(0) + *T*_*aC*_(0) + *R*_*a*_(0) + *R*_*C*_(0) + *R*_*aC*_(0) + *V*_*C*_(0) ∈ ℜ_+_. *Then*

1. 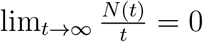
2. 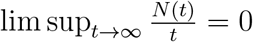
3. 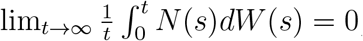, and
4. 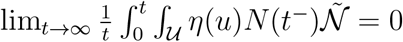, *a.s*.

#### Theorem 12.

*Let* (*S*(*t*), *I*_*C*_(*t*), *I*_*a*_(*t*), *I*_*aC*_(*t*), *I*_*c*_(*t*), *I*_*cC*_(*t*), *T*_*a*_(*t*), *T*_*C*_(*t*), *T*_*aC*_(*t*), *R*_*a*_(*t*), *R*_*C*_(*t*), *R*_*aC*_(*t*), *V*_*C*_(*t*)) *be the solution of* (3) *with initial v alues S*(0), *I*_*C*_(0), *I*_*a*_(0), *I*_*aC*_(0), *I*_*c*_(0), *I*_*cC*_(0), *T*_*a*_(0), *T*_*C*_(0),*T*_*aC*_(0), *R*_*a*_(0), *R*_*C*_(0), *R*_*aC*_(0), *V*_*C*_(0) ∈ Ω, *the co-infection disease of model* (3) *goes extinct almost surely* (lim_*t*→∞_ *I*_*aC*_(*t*) = 0) *if* 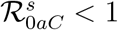.

*Proof*. Let *V*_1_ = ln *I*_*aC*_(*t*) be the Lyapunov function, then the Itô-Lévy expression of the co-infected equation of (3)

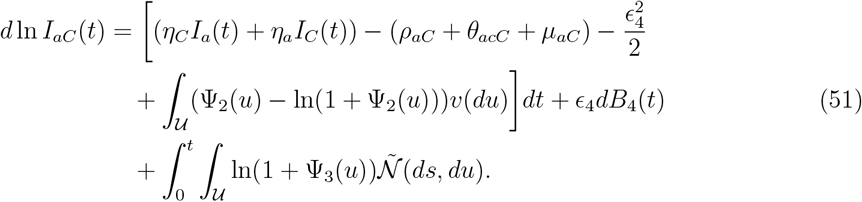

Next, we integrate both sides of (51) we obtain

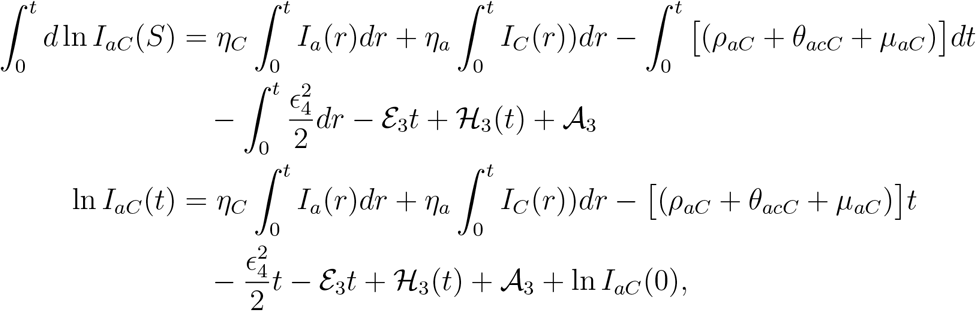

where 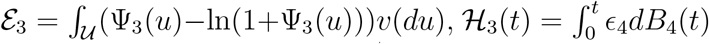, and 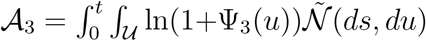.

Since 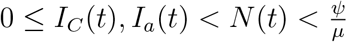, we choose a constant 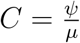, and let 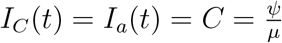 to get

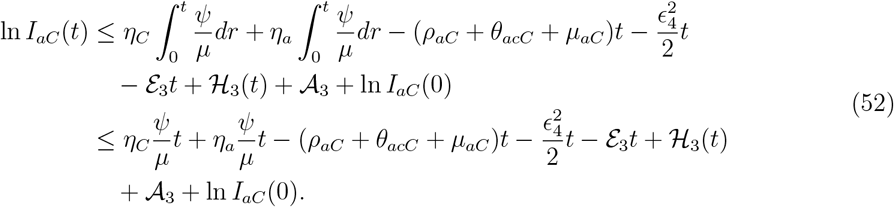

We divide through (52) by t to obtain

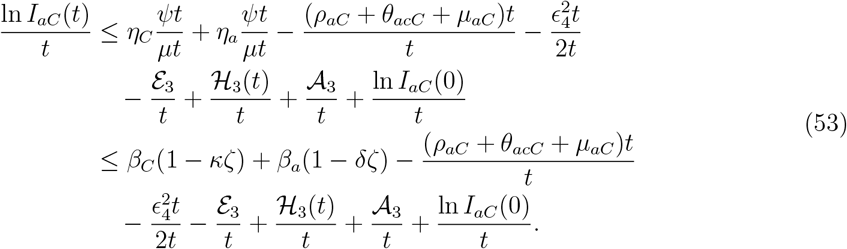

Next, we find the lim sup_*t*→∞_ on both sides, apply Lemma (1), and factorise to obtain

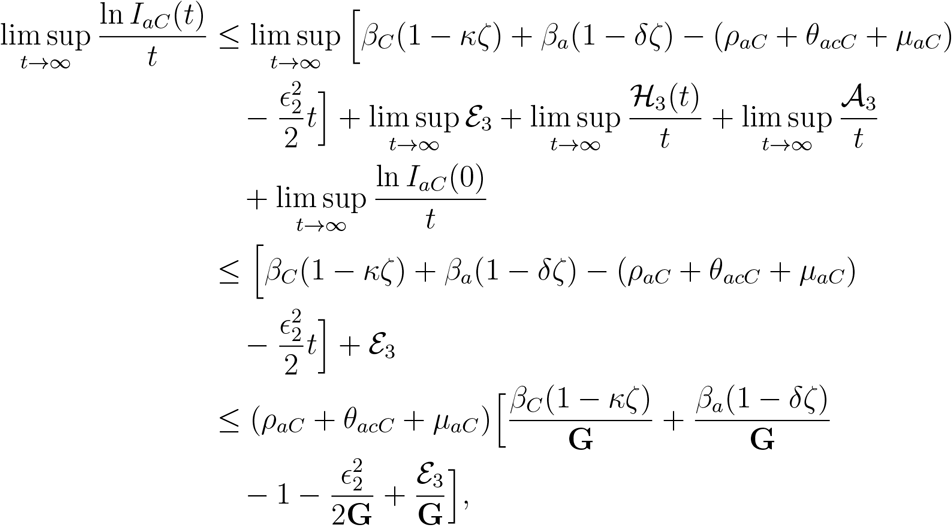

where **G** = (*ρ*_*aC*_ + *θ*_*acC*_ + *µ*_*aC*_). From (45) we have

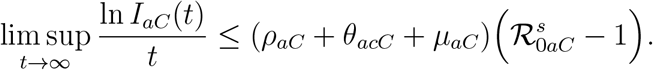

Clearly **G** = (*ρ*_*aC*_ + *θ*_*acC*_ + *µ*_*aC*_) *>* 0.

If

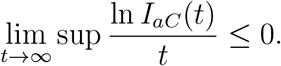

then

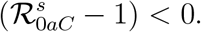

From (1) we conclude that whenever 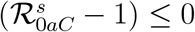,

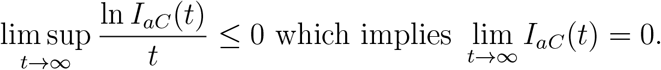

Next we determine the condition for extinction of HBV only disease in the population.

#### Theorem 13.

*Let* (*S*(*t*), *I*_*a*_(*t*), *I*_*c*_(*t*), *T*_*a*_(*t*), *R*_*a*_(*t*)) *be the solution of HBV only model from* (3) *with initial values S*(*t*), *I*_*a*_(0), *I*_*c*_(0), *T*_*a*_(0),

*R*_*a*_(0) ∈ Ω, *the HBV disease goes extinct almost surely* (lim_*t*→∞_ *I*_*a*_(*t*) = 0) *if* 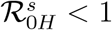.

*Proof*. Let *V*_2_ = ln *I*_*a*_(*t*) be the Lyapunov function, then by applying Itô’s formula we recall (18), which is the Itô expression for the HBV only infected class of (3)

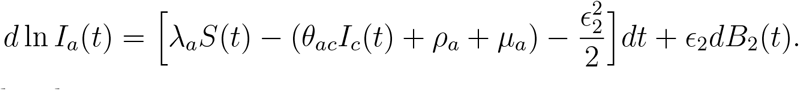

We integrate both sides to get,

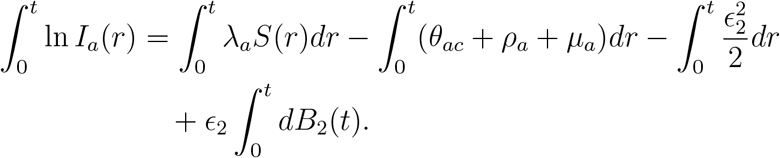

We know that 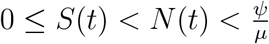, hence, we have

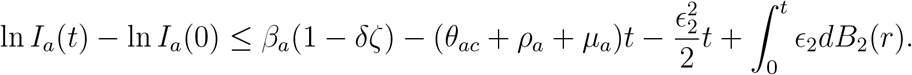

We divide through by t, find the lim sup_*t*→∞_ on both sides, apply Lemma (1), and factorise to obtain

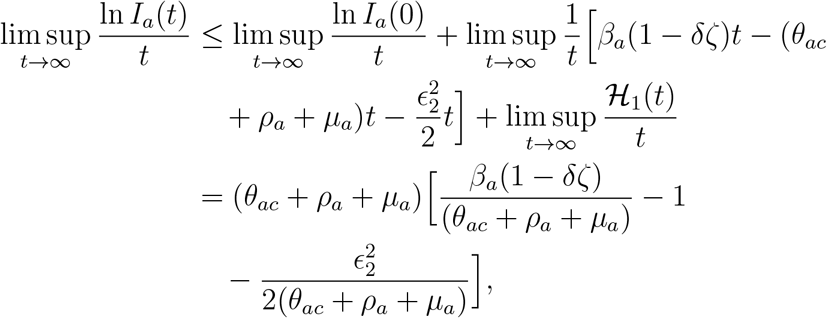

where 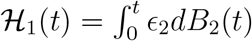, is a martingale.

From (29) we have

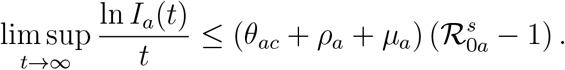

Clearly,

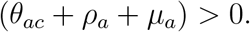

If

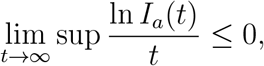

Then

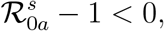

From (1) we conclude that whenever 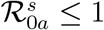,

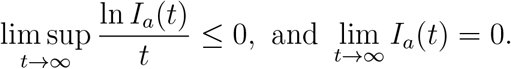

Next, we derive the condition for extinction of *I*_*C*_(*t*).

#### Theorem 14.

*Let* (*S*(*t*), *I*_*C*_(*t*), *T*_*C*_(*t*), *R*_*C*_(*t*), *V*_*C*_(*t*)) *be the solution of COVID-19 only model from* (3) *with initial values S*(0), *I*_*C*_(0), *T*_*C*_(0), *R*_*C*_(0), *V*_*C*_(0) ∈ Ω, *the COVID-19 disease goes extinct almost surely* 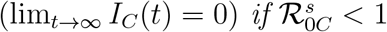.

*Proof*. Let *V*_3_ = ln *I*_*C*_(*t*) be the Lyapunov function. Applying the Itô-Lévy formula gives

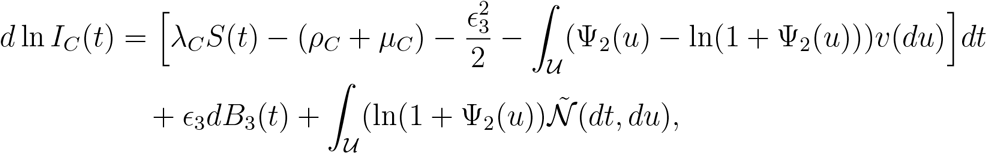

which is the Itô-Lévy expression for the COVID-19 infected class of the COVID only system of (3). Next, we integrate both sides to get,

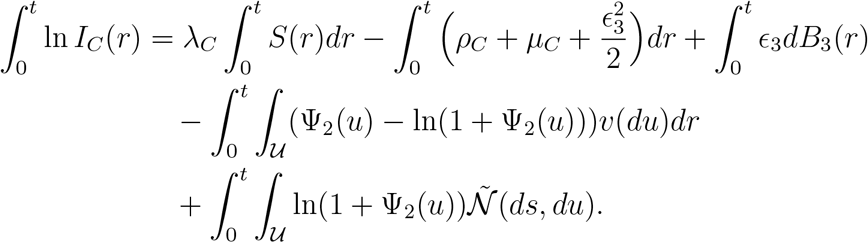

We know that 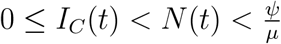, hence,

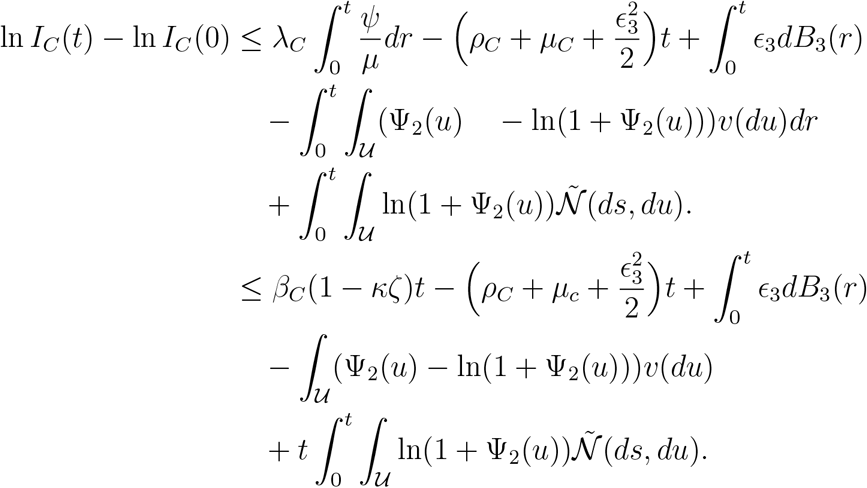

We divide through by t, find the lim sup_*t*→∞_ on both sides, apply Lemma (1), and factorise to obtain

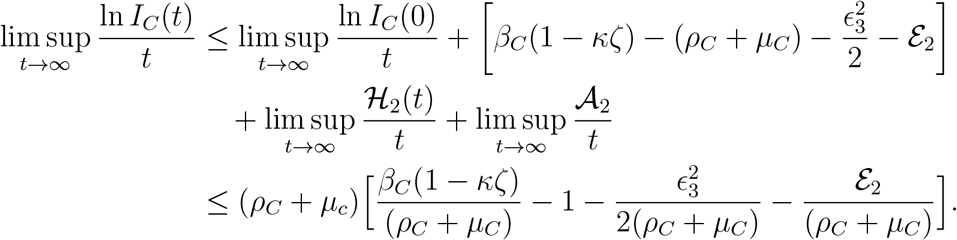

From (15) we have

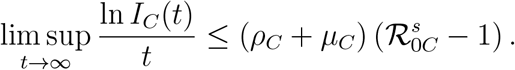

Where 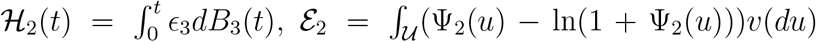, and 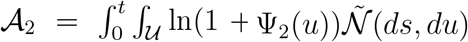 are martingales.

Clearly

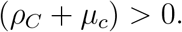

For

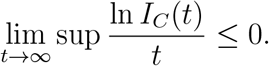

Then

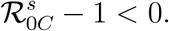

From (1) we conclude that whenever 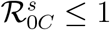,

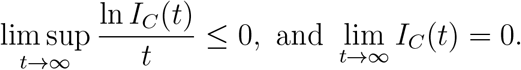

### 3.8 Persistence in Mean

Persistence of a disease means that the disease will remain endemic in the population with positive probability. We study the disease persistence for the system reported in (3) and derive that the disease persists under certain conditions.

#### Definition 4.

*[1] Persistence for Model* (3) *will hold if*,

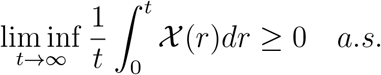

#### Lemma 2.

*[50] Set g* ∈ ℂ [0, ∞] × Ω, (0, ∞)], *assume that there exist ξ*_0_, *ξ >* 0 *such that*

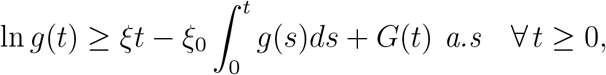

*such that G* ∈ (ℂ [0, ∞] × Ω, (0, ∞)] *satisfying* 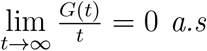. *Then* 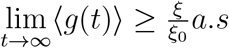.

#### Theorem 15.

*Let* (*S*(*t*), *I*_*C*_(*t*), *I*_*a*_(*t*), *I*_*aC*_(*t*), *I*_*c*_(*t*), *I*_*cC*_(*t*), *T*_*a*_(*t*), *T*_*C*_(*t*), *T*_*aC*_(*t*), *R*_*a*_(*t*), *R*_*C*_(*t*), *R*_*aC*_(*t*), *V*_*C*_(*t*)) *be a solution of system* (3) *with initial values* (*S*(0), *I*_*C*_(0), *I*_*a*_(0), *I*_*aC*_(0), *I*_*c*_(0), *I*_*cC*_(0), *T*_*a*_(0), *T*_*C*_(0), *T*_*aC*_(0), *R*_*a*_(0), *R*_*C*_(0), *R*_*aC*_(0), *V*_*C*_(0)) ∈ Ω.

1. *If* 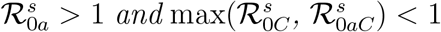, *the disease I*_*a*_ *persists in mean. In addition, I*_*a*_ *satisfies*

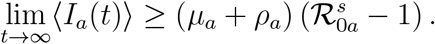
2. *If* 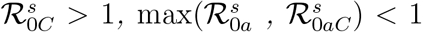, *then the disease I*_*C*_ *is persistent in mean. In addition, I*_*C*_ *satisfies*

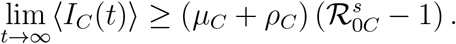

*Proof*. We begin by proving the persistence for co-infection of diseases. Thereafter, using similar techniques, we prove the persistence for singular diseases.

We recall the Itô-Lévy expression of the co-infected equation of (3).

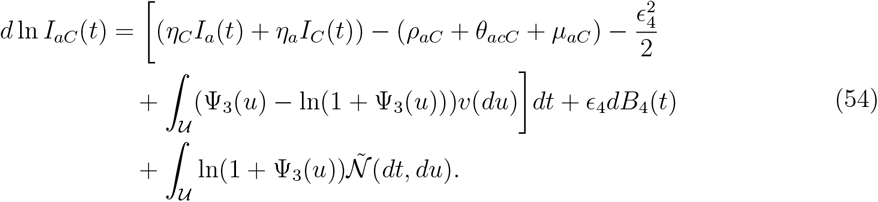

By integrating both sides of (54) we obtain

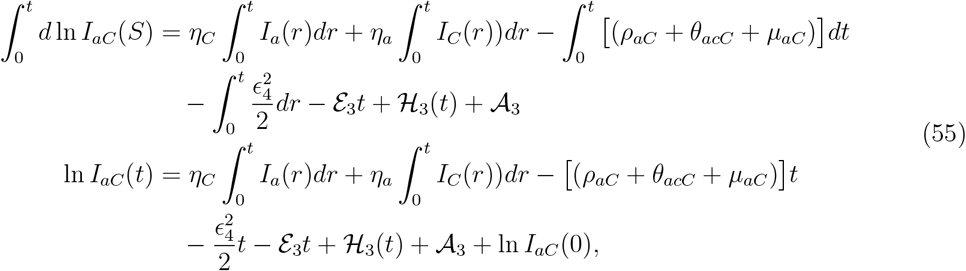

Where 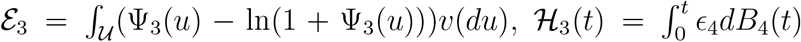, and 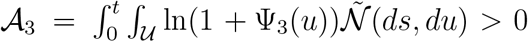. Next we divide through (55) by t, apply comparison and Definition (3) to obtain

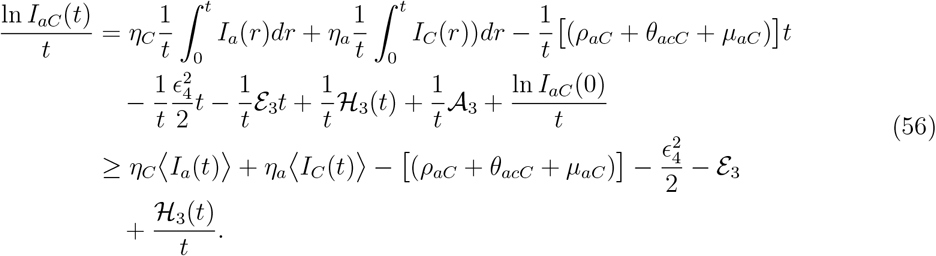

Next, we determine expressions for ⟨*I*_*a*_(*t*) ⟩, and ⟨*I*_*C*_(*t*) ⟩

By integrating the system (3) and dividing by t, to obtain the following:

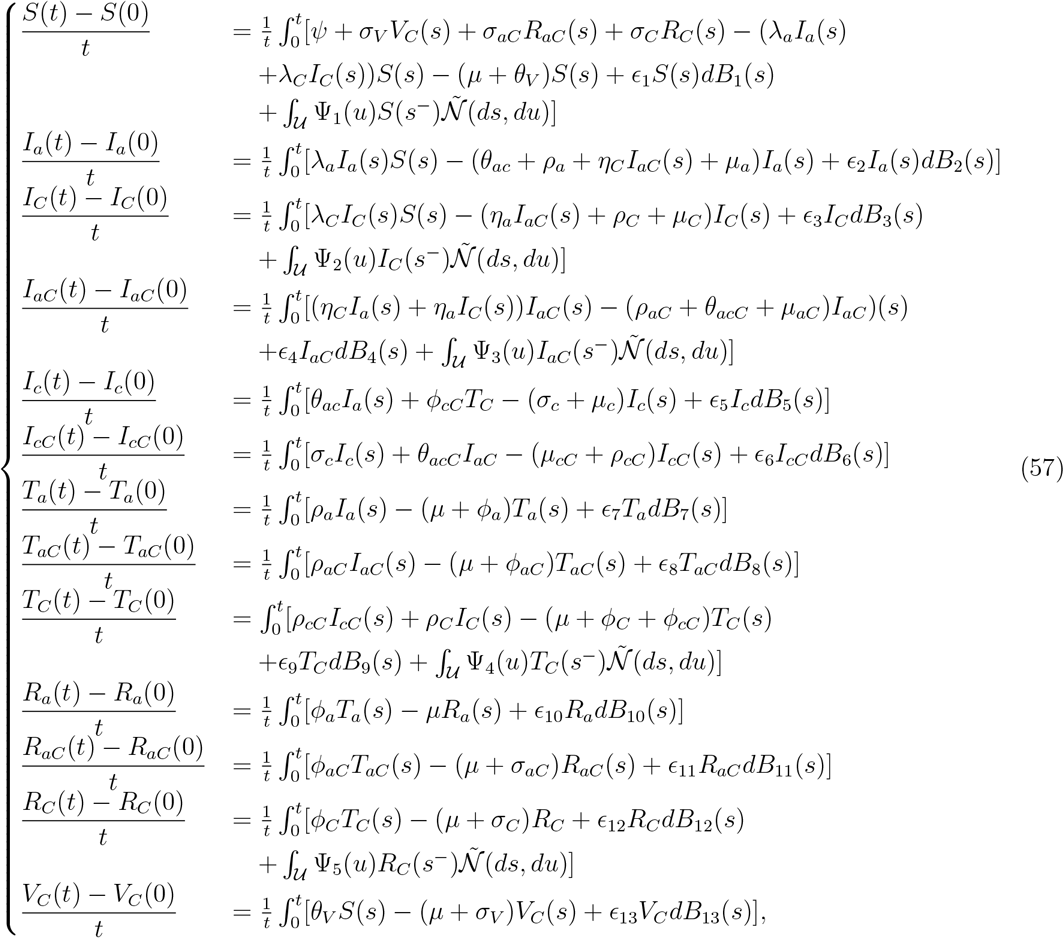

Adding terms and arrange terms of (57), rearranging and applying (3) we obtain

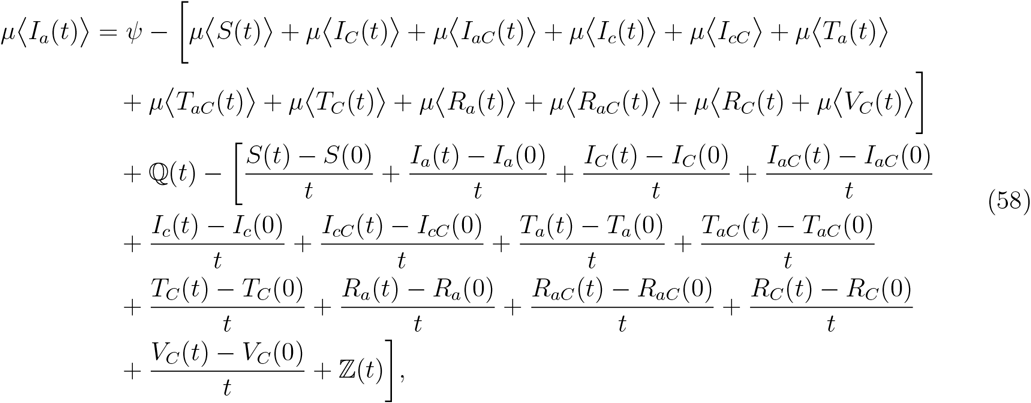

And

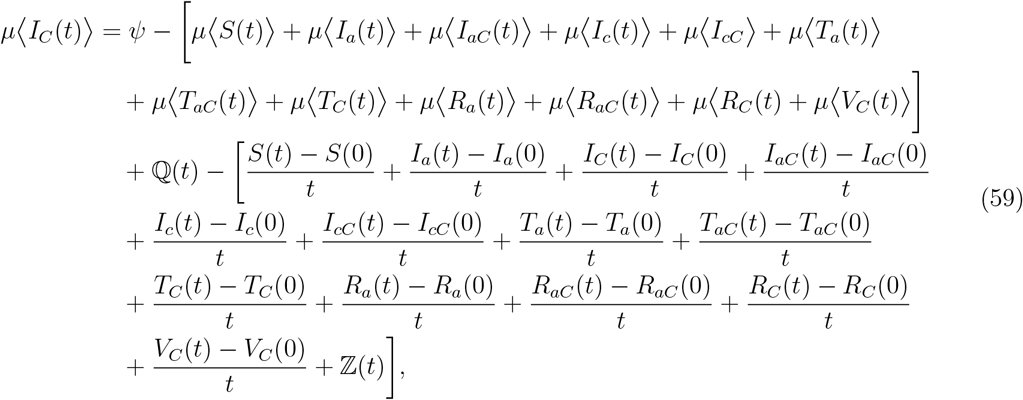

Where

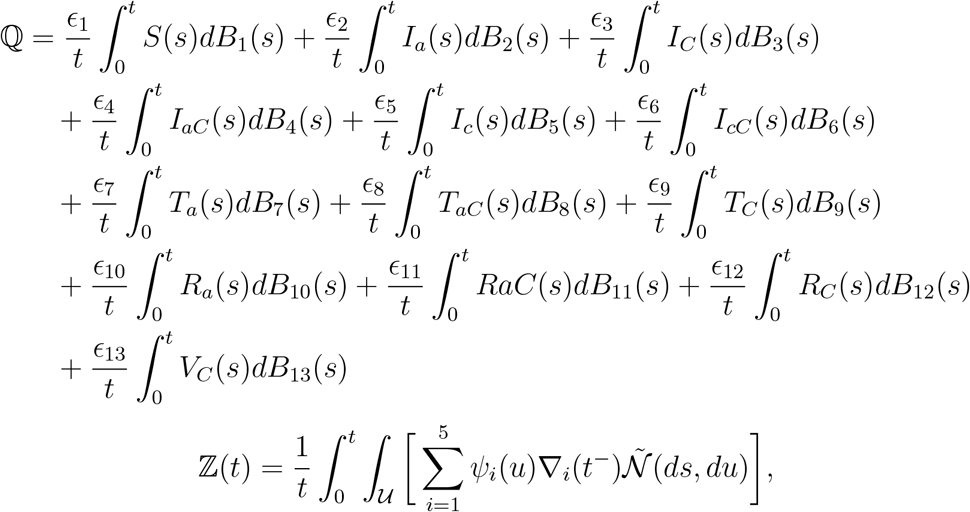

and (∇ _1_, ∇ _2_, ∇_3_, ∇_4_, ∇ _5_) = (*S, I*_*C*_, *I*_*aC*_, *T*_*C*_, *R*_*C*_). We substitute (58) and (59) into (56), take lim inf_*t*→∞_, eliminate ℤ (*t*), and apply the strong law of martingales (i.e. Lemma (1)) to get

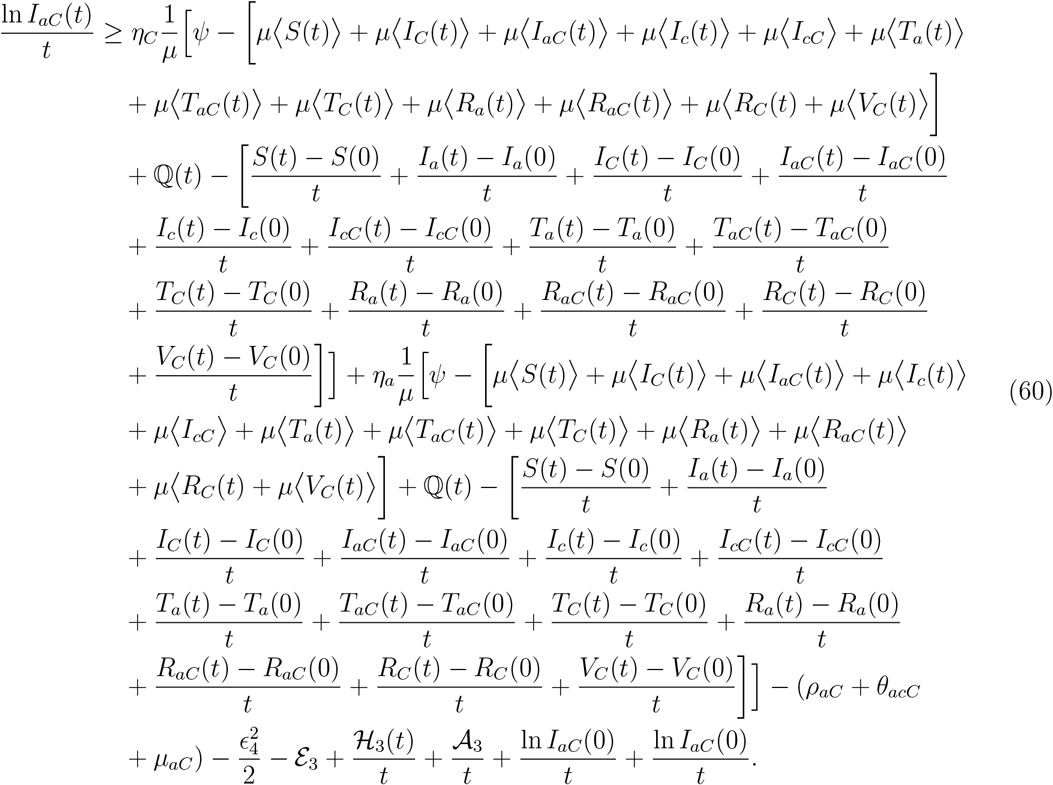

Next, find lim inf_*t*→∞_ and apply a Lemma (2) and definition (4) to obtain following results,

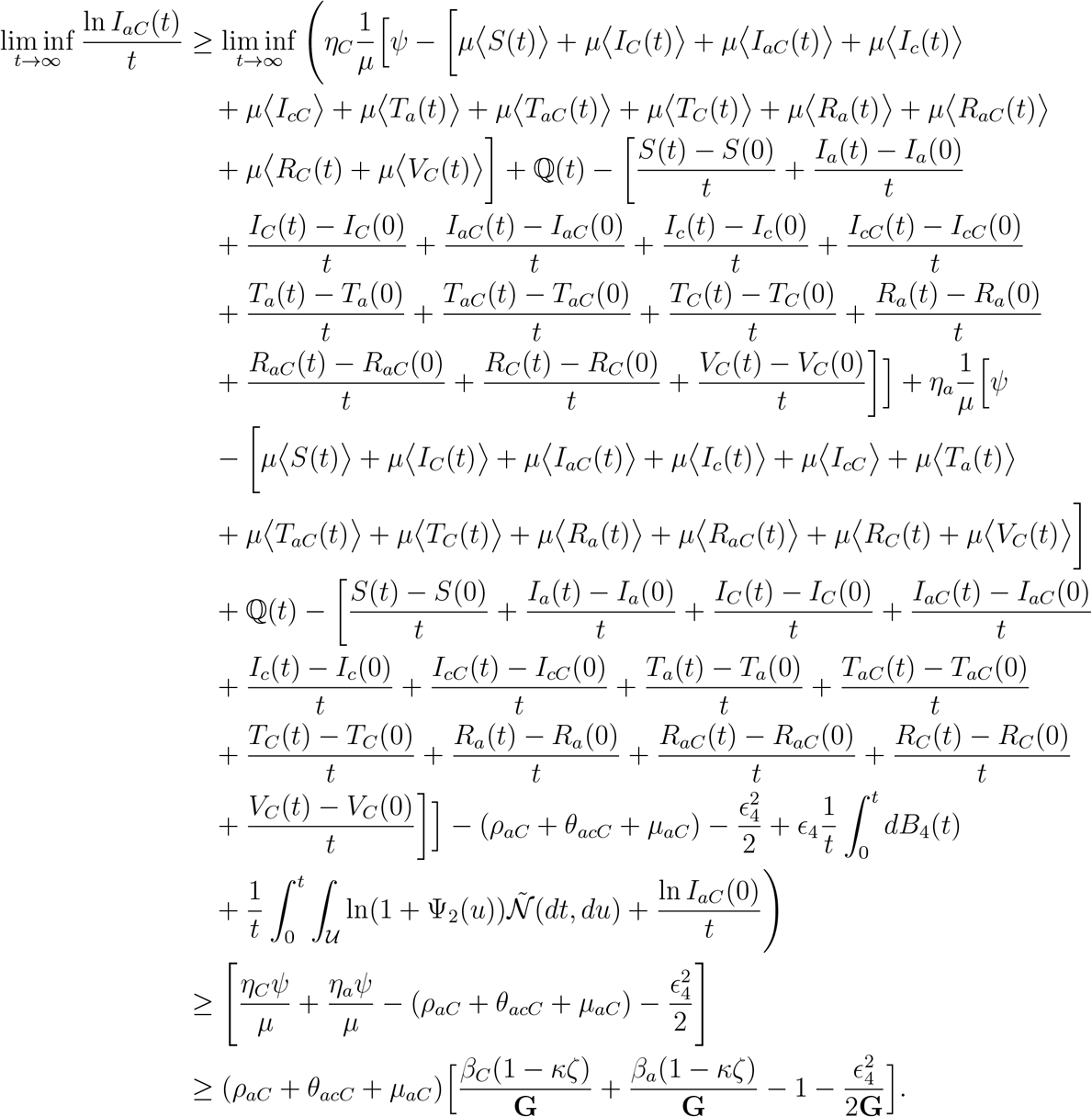

From (45) we have

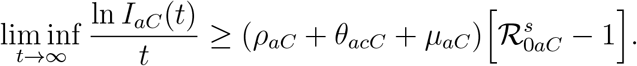

Clearly,

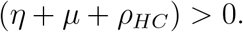

If

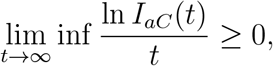

then

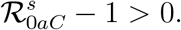

By Definition (4) Lemma (2), we conclude that whenever 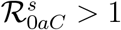,

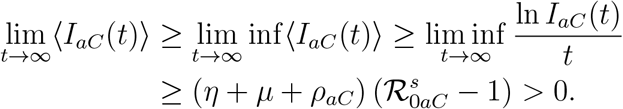

Next, we prove the persistence for HBV only Model. By rearranging terms in (59), we get

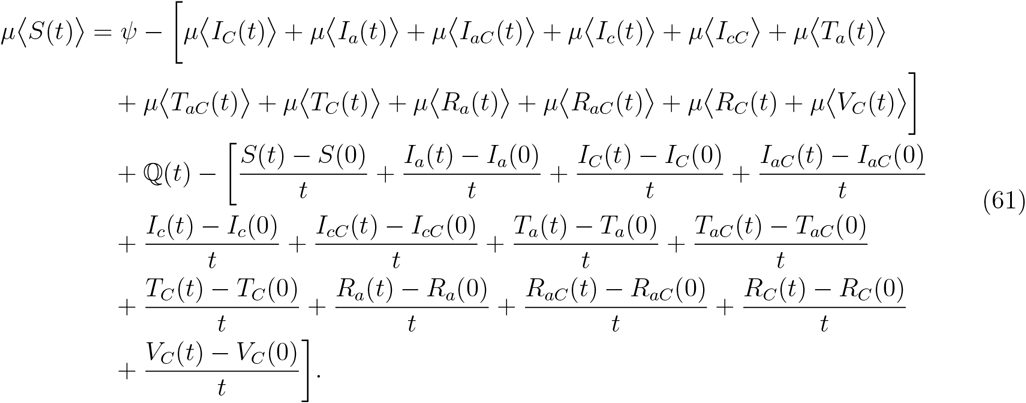

Next, we integrate both sides of (29), and eliminate positive terms to get

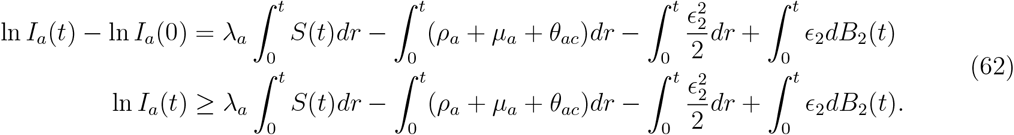

We divide (62) by t, substitute (61) into (62), to get

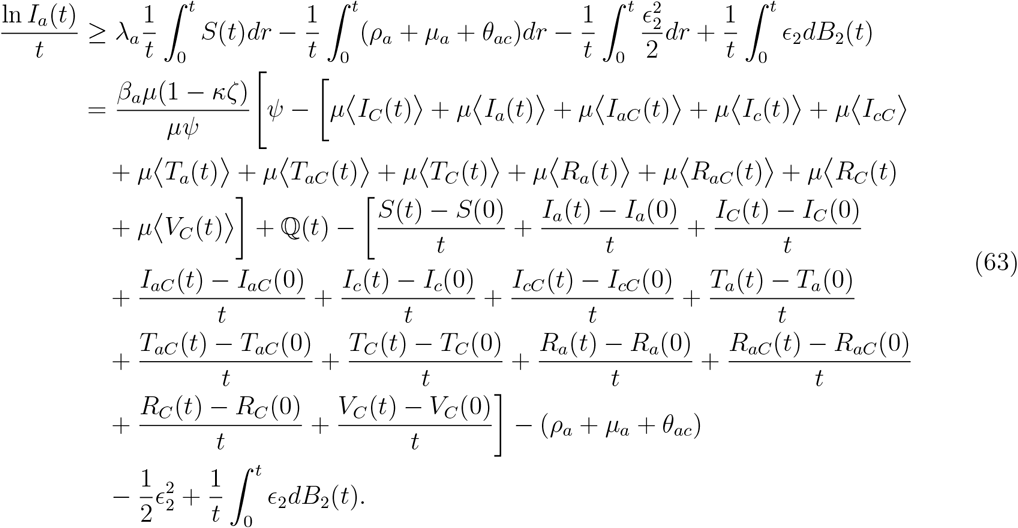

We find lim inf_*t*→∞_ and apply Definition (4) and Lemma (2) to get

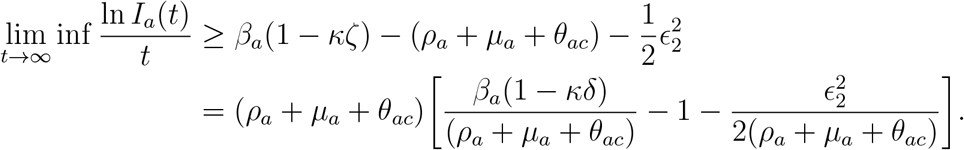

From (29) we have

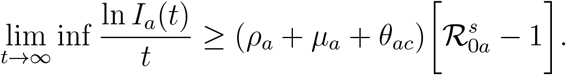

Clearly,

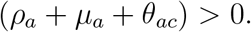

If

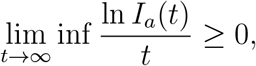

then

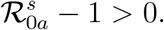

By Definition (4) and Lemma (2), we conclude that whenever 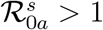,

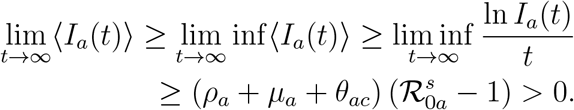

Next, we prove the persistence in mean for COVID-19. We recall the Itô-Lévy expression of the COVID-19 infected equation of (3)

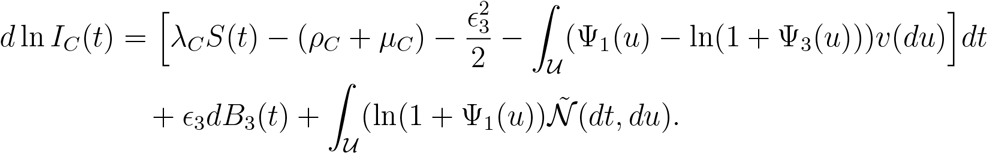

Integrating both sides we obtain

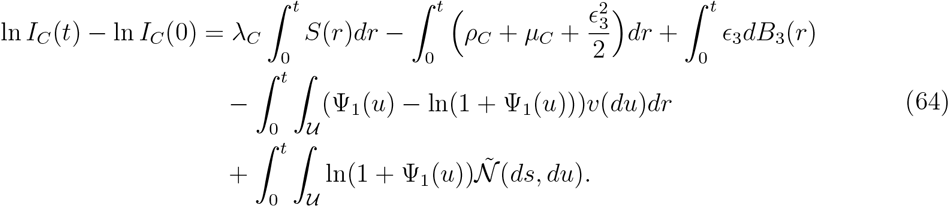

We divide (64) by t, substitute (61) into (64) to get,

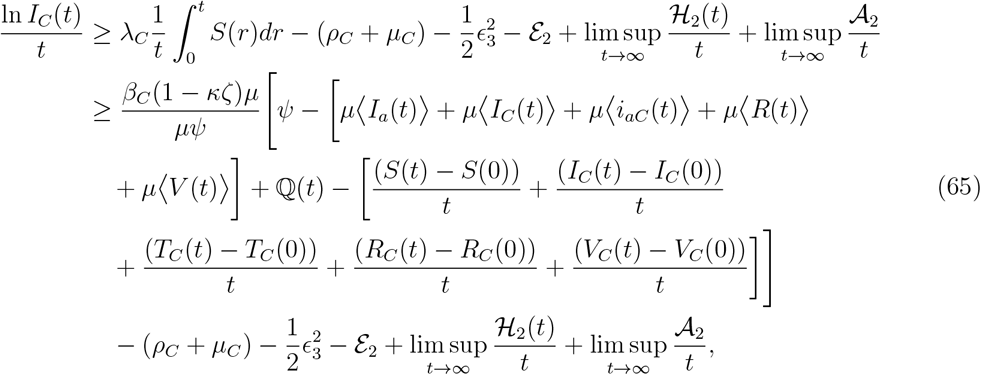

where 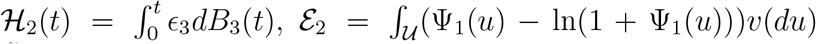, and 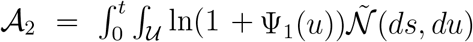 are martingales. We take lim inf_*t*→∞_ of both sides and apply Definition (4) and Lemma (2) to get

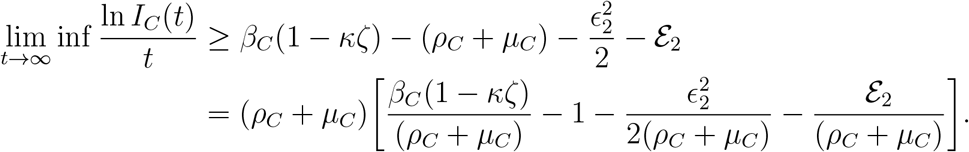

From (15) we have

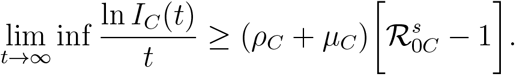

Clearly,

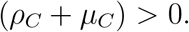

If

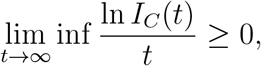

then

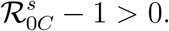

Using Definition (4) and Lemma (2), we conclude that whenever 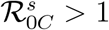,

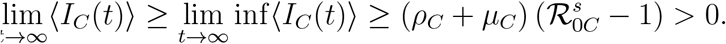

## 4 Numerical Results

In this section, we employ a standard numerical procedure to obtain numerical simulation results for the HBV-only, COVID-19-only, and HBV-COVID-19 co-infection models. The objective here is to demonstrate the theoretical results obtained earlier in this thesis.

The estimated parameters in Table (1) were derived using a combination of publicly available epidemiological data (e.g., Ghana Health Service reports, WHO reports, and peer-reviewed studies on COVID-19 and HBV), mathematical approximations using known biological and epidemiological relationships, and reasonable assumptions based on parameter values reported in similar studies when direct Ghana-specific data was unavailable.

At the beginning of the COVID-19 spread in Ghana (early 2020), the birth rate was approximately 28.60 births per 1,000 people [27]. This gives us an estimate for recruitment rate, *ψ* = 0.0286 per person.

An estimate for the natural death rate *µ* was determined using, Ghana’s life expectancy of (64.5 years) and formula

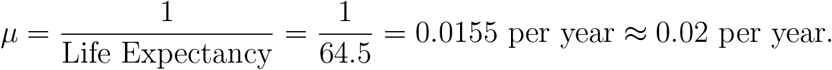

The approximation was based on published rate from other Ghanaian studies, see, e.g., [25].

COVID-19 death rate was estimated using *µ*_*C*_ = *µ* + *γ*. Available data from Ghana, shows that as of March 28, 2022 there were 1,445 COVID-19 related deaths and 161,370 confirmed COVID-19 cases [53]. Thus, the estimated COVID-19 induced death rate is

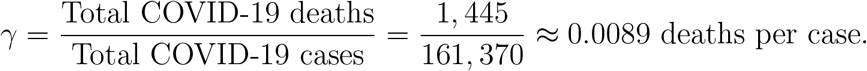

Thus, *µ*_*C*_ = *µ* + *γ* = 0.02 + 0.0089 per case.

To estimate the death rate in HBV, we first estimate the additional induced rate

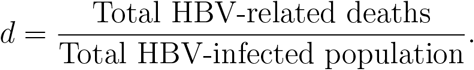

From epidemiological studies, Ghana’s HBV prevalence is estimated at 8.48% 12.30%, see, e.g., [68]. Assuming a population of 32 million, the HBV-infected population would be HBV cases = 32, 000, 000 0.011 = 3, 520, 600. According to [21], in 2022, over 15,000 people in Ghana died from hepatitis B and C-related liver diseases. Thus, the induced death rate was estimated as 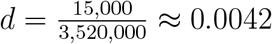. Hence the death rate in HBC state is estimated as *µ*_*a*_ = *µ* + *d* = 0.02 + 0.004 per case. With regards to COVID-19 recovery rate, it has been reported that mild cases of COVID-19 typically resolve within 10–14 days COVID-19 [12]. We estimated the recovery rate as 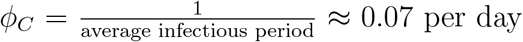. We scaled the value by 3 to reflect high recoveries in Ghana and to reflect published regional COVID-19 recovery rates, see, e.g., [17].

Acute HBV Recovery occurs over weeks to months [11]. We use and average infectious period of 60–180 days, to estimate the recovery rate as 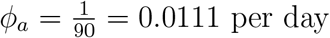.

The parameter values used along with their sources are displayed in Table 2, few values of other parameters were assumed. We sought to verify theoretical results, we selected initial values of *S*(0) = 100, *I*_*C*_(0) = 20, *I*_*a*_(0) = 20, *I*_*aC*_(0) = 5, *I*_*c*_(0) = 2, *I*_*cC*_(0) = 0, *T*_*a*_(0) = 3, *T*_*C*_(0) = 0; *T*_*aC*_(0) = 0, *R*_*a*_(0) = 2, *R*_*C*_(0) = 0, *R*_*aC*_(0) = 0, *V*_*C*_(0) = 0. The Euler-Maruyana scheme presented by [33] was employed in conducting the numerical simulations for all models. The results from numerical simulation for HBV only, COVID-19 only and confection model are presented in Figures 3 - 12.

**Table 2:** Parameters and description.

| Parameter | value | source |
| --- | --- | --- |
| $\psi$ | 0.028 births per year | [27] |
| $\theta_{ac}$ | 0.033 per day | Estimated [13] |
| $\theta_{acC}$ | 0.053 per day | Estimated [13]. |
| $\beta_C$ | [0.4800, 0.6500] per day | [14] [16] |
| $\beta_a$ | 0.4 per day | Estimated [68] |
| $\rho_a$ | 0.05 per day | Assumed |
| $\rho_{aC}$ | $\frac{1}{6}$ | Assumed |
| $\rho_C$ | $\frac{1}{6}$ | Estimated [40] |
| $\rho_{cC}$ | $\frac{1}{13}$ | Estimated [40] |
| $\phi_{aC}$ | 0.3 per day | Estimated [3] |
| $\phi_a$ | $\approx 0.05$ per day | Estimated |
| $\phi_C$ | 0.2 | [17, 7] |
| $\phi_{cC}$ | 0.03 per day | Estimated |
| $\sigma_V$ | 0.01 no unit | [35] |
| $\sigma_C$ | 0.0110 | [42] |
| $\sigma_{aC}$ | 0.002 | Assumed |
| $\theta_V$ | 0.10 | [9] |
| $\mu$ | 0.02 | [25] |
| $\mu_C$ | 0.028 per case | Estimated [25] |
| $\mu_{aC}$ | 0.03 per case | Assumed |
| $\mu_a$ | 0.021 per case | Estimated |
| $\mu_c$ | 0.024 per case | Estimated |
| $\mu_{cC}$ | 0.03 per case | Assumed |
| $\kappa$ | [0-1] | Assumed |
| $\delta$ | [0-1] | Assumed |
| $\zeta$ | [0-1] | Assumed |

**Figure 3.**
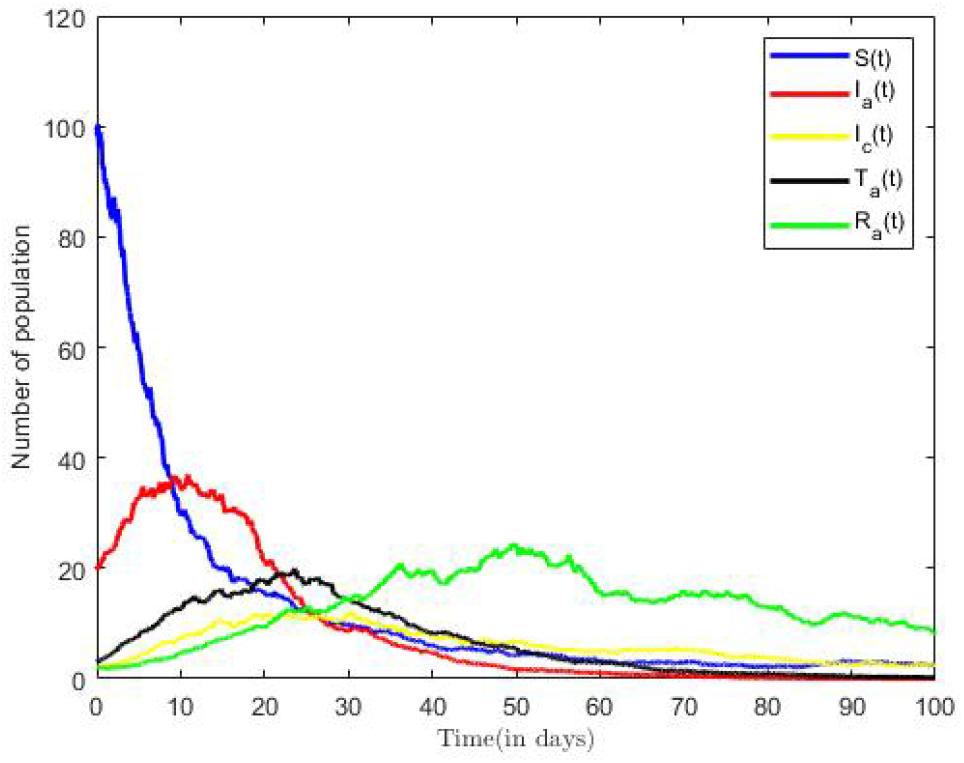
Solution for HBV Only Model (*δ* = 0.7, *ζ* = 0.6).

### 4.1 Hepatitis B Virus Only Model

From the parameter values in Table 2, we calculated the reproductive number of HBV model to be 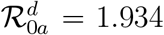. The result implies that on the average, an infected person introduced into the population can infect more than one susceptible, hence, the infection will continue. This result is consistent with other Ghanaian studies. [60] estimated the value of the Reproductive Number of HBV as ℛ_0*a*_ = 1.957, thus suggesting an epidemic population. The trajectories of susceptible, infectious, treatment and recovered classes in the HBV-only model (3) are displayed in Figure 3. Here, we fixed values of all parameters shown in Table 2.

From Figure 3, we observed that as number of infectious population increase in time, the susceptible population decrease. However, in the long run, the solution curves for both the susceptible and infective compartments behave like decreasing functions and approach a fixed point. A decrease in the number of susceptible individuals occurred rapidly within the first fifteen time steps of the epidemic, after which there was gradual decline over time. We also observe a sharp increase in the number of HBV infective within the first ten time steps and a relatively gradual increase in the number of individuals in the Treatment (or hospitalised) and other compartments within the same period. Eventually, infectious, recovered classes and other classes approach a fixed point in the long run. In other words, the results suggest that all compartments will reach their respective endemic fixed point of model (28) within a finite time and in long run. The numerical outcomes verifies that when 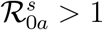, then the disease endemic equilibrium is locally and globally asymptotically stable.

The impact of various parameters on the transmission dynamics of HBV is explored. Numerical results are presented in Figure 4.We proceeded by first investigating the effects of transmission rate on population of infected individuals. This was done using selected values of transmission rates (*β*_*a*_ = 0.4, 0.3, 0.2), while all remaining parameter values of the model were held constant. From Figure 4, we observe that as values of transmission rate increases, there is an increased possibility for the population to be infected by HBV, and vice versa. Next, we investigated the effects of the transmission rate on the reproductive number of the HBV only model. Here, we held all other parameters constant, and decreased the transmission rates (*β*_*a*_ = 0.4, 0.3, 0.2). Using the varied transmission rate, we estimated Reproductive numbers as 1.934, 1.451, and 0.967 respectively. We observe from Figure 5, that for 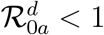, the number of infected individuals already introduced into the population declined rapidly to a zero disease free equilibrium point. The observation implies that at 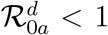, the disease will eventually become extinct. Further, it is observed from Figure 5 that an increase in the transmission rates from *β*_*a*_ = 0.3 to *β*_*a*_ = 0.4 led to increases in the Reproductive number, which translates into increases in the number of infective population over time, as well as an increase in the time to endemic equilibrium. Numerical simulations of the model in Figure 5 suggest that decreasing increasing compliance to protective measures or the decreasing disease transmission rates results in a reduction of 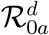, thus, reduces the average number of HBV infections in the population leading to extinction of the disease. We therefore suggest that all persons comply to preventive measures including vaccination, safe sexual practices, desist from the sharing syringes, ensuring healthcare providers use sterile, single-use needles and screen blood donations screened HBV.

**Figure 4.**
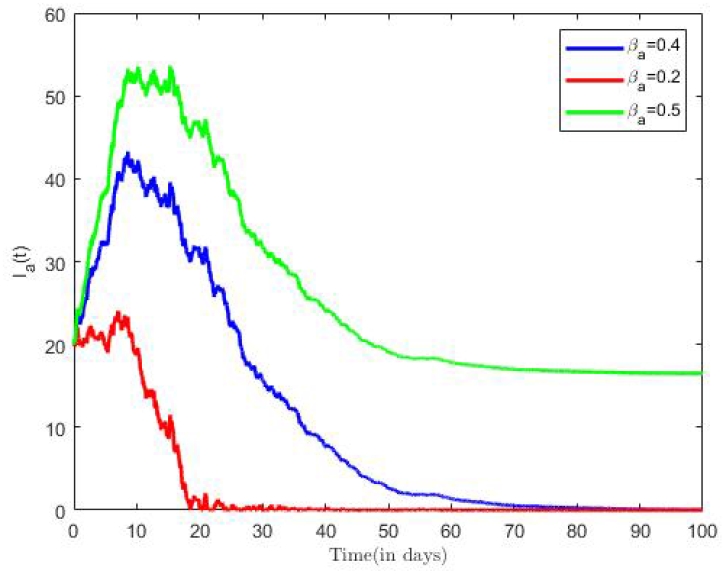
HBV Infection with varied effective contact rate.

**Figure 5.**
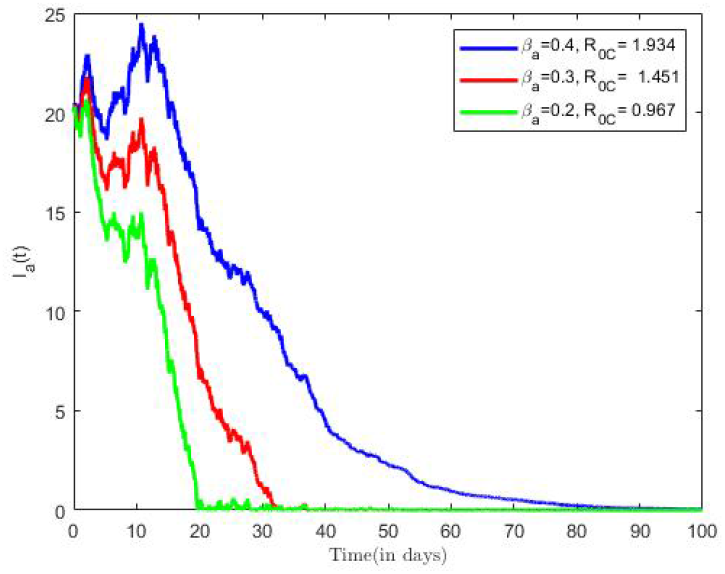
Infected population with varied transmission rate.

### 4.2 COVID-19 Only Model

From the parameter values in Table 2, we calculated the reproductive number of COVID-19 of Ghana as 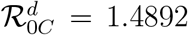, for a transmission rate on *β*_*C*_ = 0.5. The result implies that on the average, an infected person introduced into the population can infect more than one susceptible, hence, the infection will continue. This result is consistent with other Ghanaian studies, see, e.g., [54, 14]. According to [14] the estimated the value of the Reproductive number of COVID-19 in Ghana ranges between ℛ_0*C*_ = 1.47 and ℛ_0*C*_ = 2.65 for transmission rates of 0.5 and 0.9 respective, thus suggesting an epidemic population at these rates of transmission. The trajectories of susceptible, infectious, vaccinated and recovered classes in the COVID-only model (11) with only with both noise and with only Brownian noise are displayed in Figure 6 and Figure 7 respectively. Here, we fixed values of all parameters shown in Table 2.

**Figure 6.**
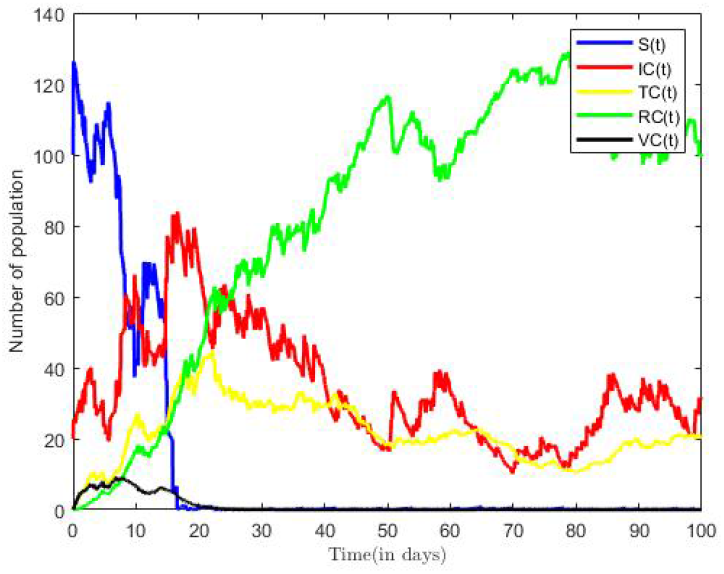
Solution to COVID-19 Only Model with both Noise (*β* = 0.5, *κ* = 0.5, *ζ* = 0.4).

**Figure 7.**
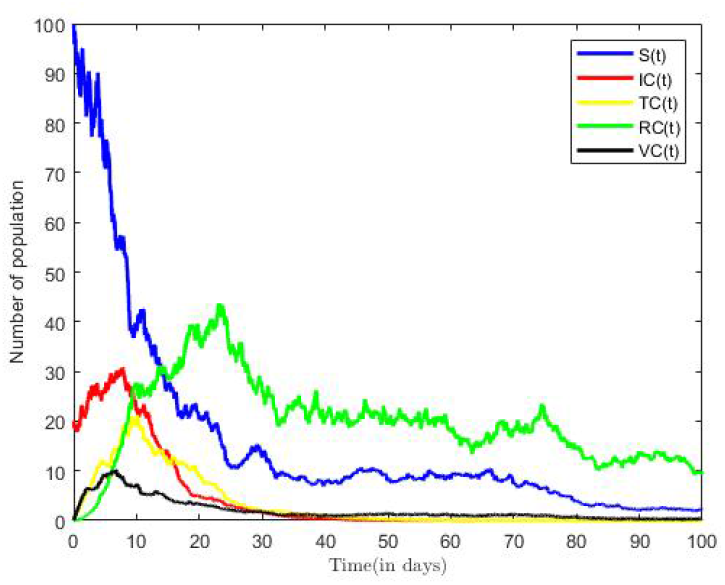
Solution to COVID-19 Only Model with Brownian noise(*β* = 0.5, *κ* = 0.5, *ζ* = 0.4).

From Figure 7, we observed that as number of infectious population increase in time, the susceptible population decrease. However, in the long run, the solution curves for both the susceptible and infective compartments behave like decreasing functions and approach a fixed point. A decrease in the number of susceptible individuals occurred rapidly within the first twenty-five time steps of the epidemic, after which there was gradual decline over time. The pattern was more rapid in Figure 6 occurring within fifteen time steps. From Figure 7, we also observe a sharp increase in the number of COVID-19 infective within the first ten time steps and a relatively gradual increase in the number of individuals in the Treatment(or hospitalised) and other compartments within the same period. Eventually, infectious, recovered, and other classes in Figure 7 approach a fixed point in the long run. In other words, the results suggest that all compartments will reach their respective endemic fixed point of model (11) within a finite time and in long run. The numerical outcomes verifies that when 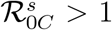, the disease endemic equilibrium is locally and globally asymptotically stable. It is worth nothing that the number of infective and time to reach endemic fixed point were greatly influenced by the inclusion of Lévy jumps in the COVID-19 only model 7. The impact of various parameters on the transmission dynamics of COVID-19 is explored. Numerical results are presented in Figure 8 and Figure 9. We proceeded by first investigating the effects of transmission rate on population of infected individuals. This was done using selected values of transmission rates (*β*_*C*_ = 0.3, 0.5, 0.6), while all remaining parameter values of the model were held constant. From Figure 8, we observe that as values of transmission rate increases, there is an increased possibility for the population to be infected by COVID-19, and vice versa.

**Figure 8.**
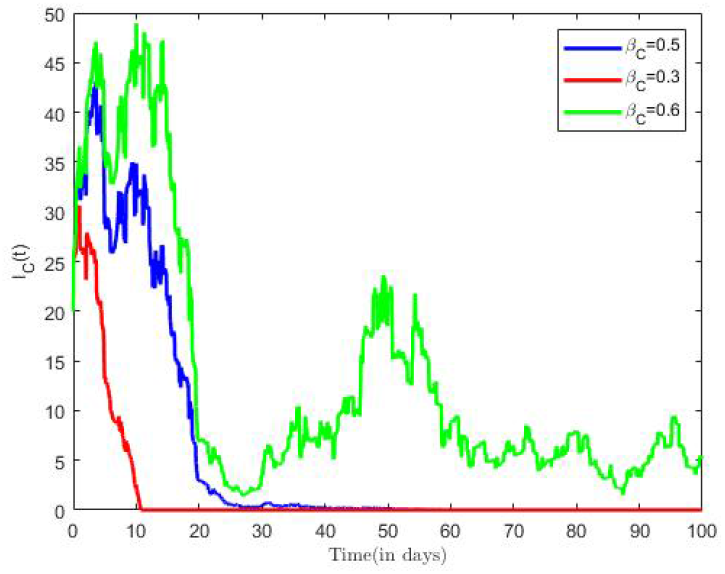
Solution to COVID-19 Only Model with Both Noise (*β* = 0.5, *κ* = 0.5, *ζ* = 0.4.

**Figure 9.**
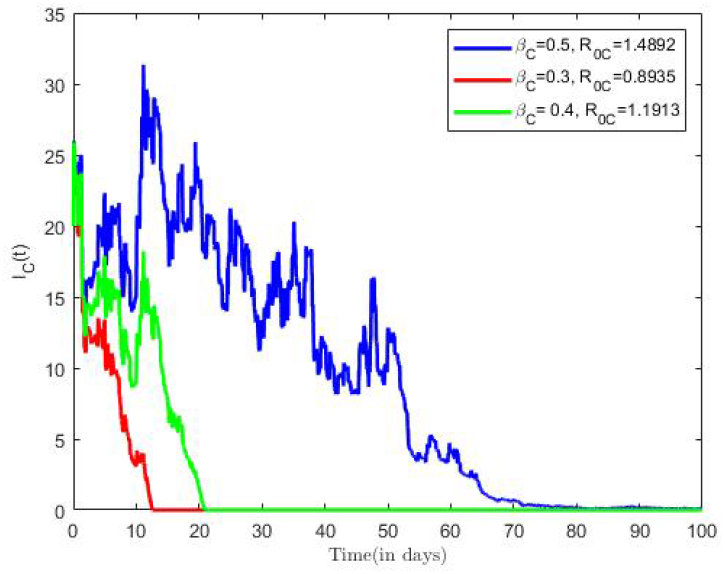
Solution to COVID-19 Only Model with Both Noise (*β* = 0.5, *κ* = 0.5, *ζ* = 0.4).

Next, we investigated the effects of the transmission rate on the reproductive number of the COVID-19 only model. Here, we held all other parameters constant, and decreased the transmission rates *β*_*C*_ = (0.3, 0.5, 0.6). Using the varied transmission rates, we estimated Reproductive numbers as 0.8935, 1.1913, and 1.4894 respectively. We observe from Figure 5, that for 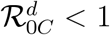, the number of infected individuals already introduced into the population declined rapidly to a disease free equilibrium point.

The observation from 8 establishes that fact that at 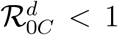, COVID-19 diseases becomes extinct as it approach a zero endemic point. Further, it is observed from Figure 8 that an increase in the transmission rates from *β*_*C*_ = 0.3 to *β*_*C*_ = 0.6 led to increases in the Reproductive number, which translates into increases in the number of infective population over time, as well as an increase in the time to endemic equilibrium. Numerical simulations of the model in Figure 8 also suggest that increasing compliance to protective measures *κ* or the decreasing disease transmission rates results in a reduction of 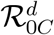, thus, reduces the average number of COVID-19 infections in the population leading to extinction of the disease. We therefore suggest that all persons comply to preventive measures including social distancing, wearing facial mask, and the use of alcohol based sanitizer.

### 4.3 Co-infection Model

From the parameter values in Table 2, we calculated the reproductive number of co-infection model to be 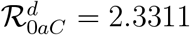 for (*β*_*C*_ = 0.5, *β*_*a*_ = 0.4). The result implies that on the average, an infected person introduced into the population can infect more than two susceptible, hence, the infection will continue. The trajectories of susceptible, infectious, treatment, recovered classes and other compartments in the co-infection model (3) are displayed in Figure 10. Here, we fixed values of all parameters shown in Table 2.

**Figure 10.**
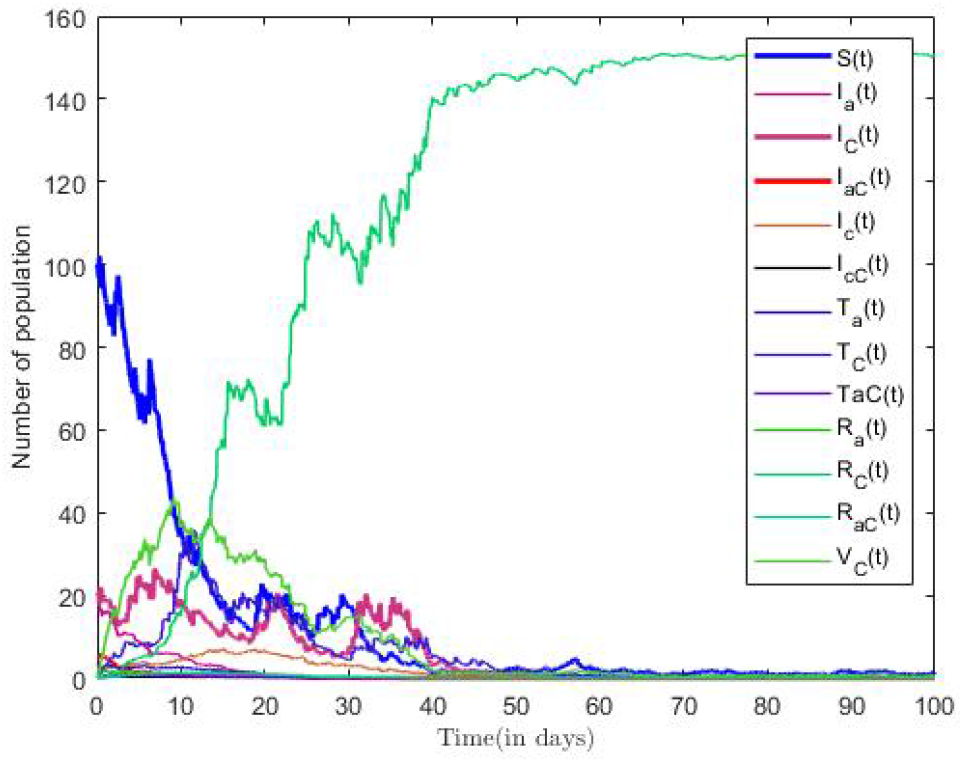
Solution for Co-infection Model (*κ* = 0.5, *δ* = 0.7, *ζ* = 0.6).

From Figure 10, we observe that as number of co-infectious population increase in time, the susceptible population decrease. However, in the long run, the solution curves for both the susceptible and infective compartments behave like decreasing functions and approach a fixed point. A decrease in the number of susceptible individuals occurred rapidly within the first fifteen time steps of the epidemic, after which there was gradual decline over time. We also increase in the number infected, Treatment and other compartments over the period. Eventually, infectious, recovered classes and other classes approach a fixed point in the long run. In other words, the results suggest that all compartments will reach their respective endemic fixed point of model (3) within a finite time and in long run. The numerical outcomes verifies that when 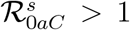, then the disease endemic equilibrium is locally and globally asymptotically stable.

The impact of various parameters on the transmission dynamics of HBV is explored. Numerical results are presented in Figure 11 and Figure 12. We proceeded by first investigating the effects of transmission rate on population of infected individuals. This was done using varied pairs of values of transmission rates (*β*_*a*_ = 0.2, 0.3, 0.4), and (*β*_*C*_ = 0.3, 0.4, 0.5), while all remaining parameter values of the model were held constant. From Figure 11, we observe that as values of transmission rate increases, there is an increased possibility for the population to be co-infected with HBV, and COVID-19. Further, we investigated the effects of the transmission rate on the reproductive number of the co-infection model using the aforementioned rates. The estimated Reproductive numbers as 1.3057, 1.8184, and 2.3311 respectively. It is observed from Figure 11 that an increase in the transmission rates, *β*_*a*_, and *β*_*C*_, resulted in increases in the Reproductive number, which translates into increases in the number of infective population over time, as well as an increase in the time to endemic equilibrium.

**Figure 11.**
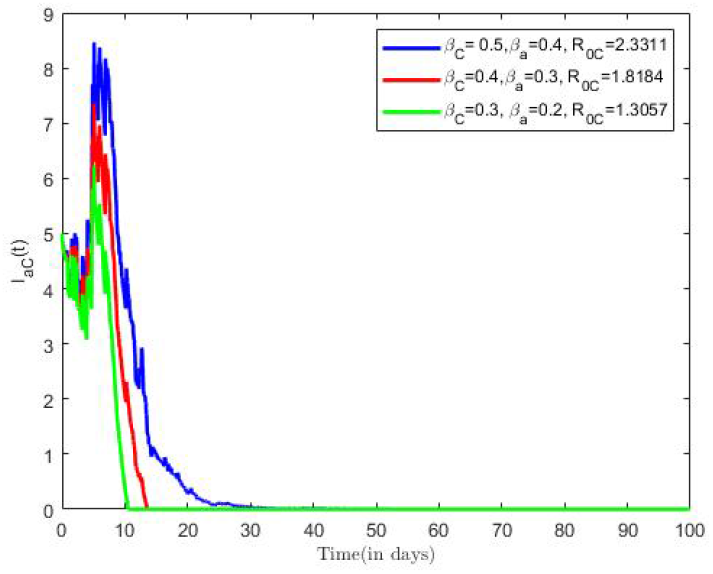
Co-Infection with varied effective contact rate and *δ* = 0.7, *κ* = 0.5, *ζ* = 0.6.

**Figure 12.**
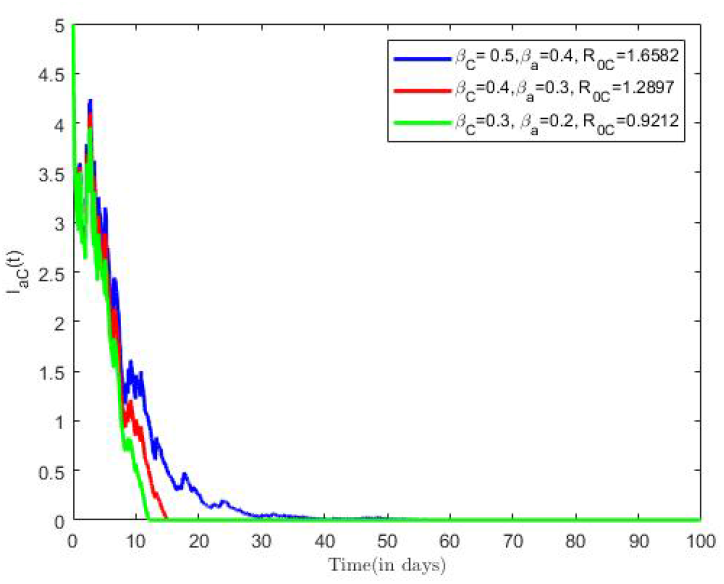
Co-infected population with varied transmission rate and *δ* = 0.9, *κ* = 0.9, *ζ* = 0.6.

Next, we investigated the effects of the compliance with preventive measures on the reproductive number of the Co-infection model. Here, we increased *κ* from 0.7 to 0.9, *δ* from 0.7 to 0.9, all other parameters were held constant, which earlier paired varied transmission rates (*β*_*a*_ = 0.2, 0.3, 0.4), and (*β*_*C*_ = 0.3, 0.4, 0.5) were utilised. We estimated Reproductive numbers as 0.9212, 1.2897 and 1.6582 respectively. It is observed from Figure 12 that an increase both COVID-19 and HBV compliance rate resulted in a reduction of the Reproductive number, which translates into decreases in the number of co-infective population over time as compared to Figure 11.

Numerical simulations of the model in Figure 12 also suggest that increasing compliance to COVID-19 and HBV preventive measures will results in a reduction of number of co-infected individuals in the population leading to extinction of the disease. We therefore suggest that all persons comply to preventive measures including social distancing, wearing facial mask, and the use of alcohol based sanitizer, HBV vaccination, safe sexual practices, desist from the sharing syringes, and ensuring healthcare providers use sterile.

## 5 Conclusion

The majority of issues in the real world are not deterministic. Because of its proximity to ambient sounds, the stochastic models allow us to model epidemic diseases in a more realistic manner.

In this paper, two distinct diseases were used to study a stochastically perturbed SITRVS-type model. An emphasis was placed on explaining the effect of small fluctuations and perturbations due to sudden environmental shocks in the transmission dynamics of the co-infection, hence the inclusion of Poisson-driven Lévy noise. To determine the conditions for the local asymptotic stability of the equilibria, we first computed the reproductive number and the equilibria for the underlying stochastic model (3). Furthermore, we investigated the conditions of extinction and persistence of HBV only, COVID-19 only, and co-infection models. We observe that persistence for all sub-models and co-infection model were dependent on the intensity of the Brownian noise and the values of epidemic parameters involved in disease transmission.

Additionally, we demonstrated the influence of different parameters on the infectious compartments by presenting several numerical simulation results using the Euler-Murayama scheme.

## Data Availability

All data produced in the present work are contained in the manuscript

## 6 Declarations

### 6.1 Availability of data and materials

The authors confirm that the data used in this study are available within the article.

### 6.2 Competing Interest

The authors declare that they have no competing interests.

### 6.3 Funding

This research did not receive any funding.

### 6.4 Authors’ Contribution Statement

**MAP** made the main contribution to this manuscript including theoretical formulation, analysis and simulations. **SEM** supplied several suggestions in theoretical part, editing and proofreading the manuscript. **SMN** supplied several suggestions in theoretical part, editing and proofreading the manuscript. All authors read and approved the final manuscript.

## Notes

### Competing Interest Statement

The authors have declared no competing interest.

### Funding Statement

This study did not receive any funding

### Summary of Updates

The title of paper has been modified. Authors have been updated, Abstract has been updated. Section 1, has been updated, Section 2 has been updated. Section 3, Section 4, Section 5 have been revised. Reference section has also been updated.

